# The Impact of Rural Hospital Closures and Mergers on Health System Ecologies: A Scoping Review

**DOI:** 10.1101/2025.01.31.25321470

**Authors:** Alison Coates, Janice Probst, Kanika Sarwal, Suhaib Riaz, Agnes Grudniewicz

## Abstract

Despite playing a pivotal role in rural community health services delivery and in local economies, rural hospitals in the United States have closed or merged with larger health networks at alarming rates. This scoping review examines what is known about the impacts of rural hospital closures and mergers, synthesizing the literature across 2010-2024. Most of the identified studies examined closures, primarily reporting on community impacts. Using this literature, we inductively derived a new Health System Ecologies Impact Matrix research tool to assess knowledge related to health system changes. Knowledge gaps remain related to financial, workforce, and utilization-related outcomes, and little is known about closure impacts on neighboring hospitals and communities. Few studies report effects of rural hospital mergers, with those analyses primarily focusing on financial and utilization outcomes for the merged hospital. No studies examined the impacts of rural hospital mergers on patients or individuals and their social environments.

**Protocol:** The registered scoping review protocol may be accessed on OSF: https://doi.org/10.17605/OSF.IO/GEKJC

## Introduction

Rural hospitals have long been a keystone for delivering health services, participating in population health initiatives, sustaining healthy rural communities, and contributing to the economic health of their regions. Financial precarity among rural hospitals has increased, especially in states that did not expand Medicaid as provided for by the Affordable Care Act (ACA) of 2010 (Bai et al., 2020). Across the country, many rural hospitals have closed leaving rural communities with diminished access to care, or have sought merger or affiliation models to remain viable. Given that rural communities experience multiple disparities in health outcomes and social determinants of health compared to the United States (US) population (*CMS Framework for Advancing Health Care in Rural, Tribal, and Geographically Isolated Communities*, n.d.), changes in availability of hospitals and health services could exacerbate rural health inequity.

Approximately half of the hospitals in the US are located in rural areas, with 22% of these located in isolated regions, far from other population centers (Freeman et al., 2015). Most rural hospitals are small; 88% have fewer than 100 beds, and 56% have fewer than 26 beds (Freeman et al., 2015). Unsurprisingly, rural hospitals average lower volumes than their urban counterparts (Harrington et al., 2020). In addition to traditional inpatient, emergency, obstetric, and surgical services, rural hospitals in the US often provide outpatient primary and specialty care, diagnostic services, preventive health services, and participate actively in population health initiatives (Freeman et al., 2015).

Beyond their central role in health services delivery, rural hospitals contribute substantially to their local economies (Cordes et al., 1999; Frakt, 2019; McDermott et al., 1991; Wishner et al., 2016). These hospitals are usually among the largest employers in their region (Murphy et al., 2018; D. Williams et al., 2020), providing jobs across a variety of skill and income levels. In addition to health care providers of various types, hospitals employ clerical and technical staff to support scheduling, billing, and maintenance needs. Hospitals are thus both sources of income and employer-based health insurance for rural regions. Rural hospitals also sustain employment in other industries that support the hospital such as food, laundry, and construction (Frakt, 2019). All of these working individuals spend money in their local communities, contributing to a sustainable economy (Frakt, 2019; Holmes et al., 2006). Access to health care provided by rural hospitals impacts the broader community economy by attracting businesses and industry investments to the region and improving businesses’ ability to recruit employees (McDermott et al., 1991). While playing a central role in rural community well-being, the hospital must overcome a number of challenges (Douthit et al., 2015): population-related (Cosgrove, 2018; Coughlin et al., 2019; Matthews et al., 2017; Meit et al., 2014), geographic (Harrington et al., 2020; Lam et al., 2018), workforce-related (Coates et al., 2021; Cosgrove, 2018; Kaufman et al., 2016; US Department of Health and Human Services et al., 2020), and structural (Carroll et al., 2022; Probst et al., 2019). Several studies suggest that lower profitability (Cosgrove, 2018; Kaufman et al., 2016), low patient volume (Kaufman et al., 2016), and smaller market shares are antecedents of hospital closure (Kaufman et al., 2016). Financial distress is also reported to contribute to mergers or affiliations (Noles et al., 2015) as rural hospitals look to improve financial performance in order to survive (D. Williams et al., 2020).

The incidence of rural hospital closures in the US has been on the rise for more than a decade (Cosgrove, 2018; Sheps Center, n.d.). Recognizing the challenges of providing hospital services in rural areas under conventional payment models, Medicare designed reimbursement alternatives including the Critical Access Hospitals (CAH), Sole Community Hospitals (SCH), Medicare Dependent Hospitals (MDH), and Rural Referral Centers (RRC) designations (Freeman et al., 2015). Despite these alternative payment mechanisms, 146 rural hospitals closed between 2010 and 2023, hitting an annual peak of 18 closures in 2020 (Sheps Center, n.d.). Almost 60% of these closures resulted in the loss of a geographically isolated hospital designated as CAH, SCH, MDH, or RRC (Sheps Center, n.d.). Rural hospital closures are linked to impacts on access to and quality of care, patient health outcomes, effects on rural economies, and changes in determinants of health (Carroll et al., 2022).

In addition to the increase in rural hospital closures, the 2000s have seen a strong trend towards health system consolidation through mergers and acquisitions (Oyeka et al., 2018). From 2010 to 2016, the US averaged 44 rural hospital mergers per year, representing a 200% annual increase over the prior 5-year period (D. Williams et al., 2020). Mergers or affiliations may allow rural hospitals to achieve efficiencies of scale that they cannot as small, standalone entities (D. Williams et al., 2020). Rural hospitals were more likely to merge if they had lower profit margins, less capital available to cover debts, and older infrastructure and equipment required to operate and maintain the facility (D. Williams et al., 2020). Recently, researchers have raised concerns that increased market power of hospital networks could lead to higher health care prices and that the acquiring health systems might consolidate service lines to reduce duplication, leading to a loss of services in rural communities (Carroll et al., 2022). The literature on rural hospital mergers to date affords few explorations of these or other outcomes.

Given the role rural hospitals play in the health and well-being of their communities, closures and mergers may have broad and deep impacts. In this scoping review, we explore what is known about the impacts of rural hospital closures and mergers. We examine the recent literature on rural hospital closures and mergers in the US to understand what we know and do not know about who or what is affected and the outcomes they experience.

### New Contribution

In this scoping review, we use the literature about the effects of rural hospital closure and mergers to derive a novel framework for understanding impacts of health system changes. We derive a new research tool: the Health System Ecologies Impact Matrix. We map the identified literature onto this Impact Matrix to summarize what we know about the effects of closures and mergers of rural hospitals, and to highlight important knowledge gaps. To our knowledge, no prior review has taken a systematic approach to understanding the range of potential impacts of health system changes and measured the completeness of our knowledge across that spectrum.

## Methods

### Selection of data sources

We identified literature for this study using a scoping review approach using Joanna Briggs Institute methodology (Peters et al., 2020); a protocol was registered on Open Science Framework (https://doi.org/10.17605/OSF.IO/GEKJC). In consultation with an academic librarian, we developed a comprehensive search strategy to incorporate appropriate key terms, synonyms, and related subject headings pertaining to two main concepts: *rural hospitals* and *closures or mergers*. We executed the search in Medline, Scopus, and Business Source Complete databases, in addition to searching a comprehensive set of gray literature sources. Articles were included if they 1) discussed any outcomes or impacts of hospital closures and/or mergers, 2) focused on rural hospitals or outcomes for rural hospitals were reported separately from urban hospitals. Both title and abstract screening and full-text screening were conducted independently by two of the authors. For the subset of included records which reported the results of research studies related to closure or merger outcomes (“primary studies”), we extracted information about the study objectives, methods, design, data sources, and outcomes. For full details of the search, screening, and extraction processes, see Supplemental Material.

### Analysis of the literature

We approached the review of the literature in two phases. First, we used the included literature to inductively derive an analytical framework of outcomes and affected entities. In the second phase, we applied our novel framework to the included literature to describe the body of knowledge: we describe what we do and do not know about the impacts of rural hospital closures and mergers. We describe both phases of analysis below.

#### Phase 1: Developing an analytical framework

We searched for existing frameworks to categorize our outcome data; however, none captured the full breadth and scope of outcomes or the diversity of affected entities which emerged during our data extraction. Many health care models (Coburn et al., 2022; Donabedian, 2005; Ferlie & Shortell, 2001; Levesque et al., 2013) focus on the institutions being studied and their patients, with little to no inclusion of other entities. Because rural hospitals play broad roles in their communities beyond the provision of health services, models that narrowly apply to health-related factors or entities were insufficient for our purposes.

The social ecological model considers four levels of factors impacting health: individual, relationships, community, and society (*The Social-Ecological Model*, 2022). This model, which has been widely applied and adapted in research and practice in health promotion and related fields (Golden & Earp, 2012; McCloskey et al., n.d.; Richard et al., 2011), provided a starting point for our classification of affected entities. While the social ecological model did not fully capture the variety in affected entities, it offered a recognizable framework upon which to build.

##### Categorizing impacts reported in primary studies

We reviewed the extracted outcomes reported in the included studies and labelled them with codes that captured the individual outcomes measured (*what*) and *who* or *what* was reported to experience the outcome. These outcome measures were grouped into five categories: financial, well-being, workforce, utilization, and access to care (Figure 1).

**Figure 1:**
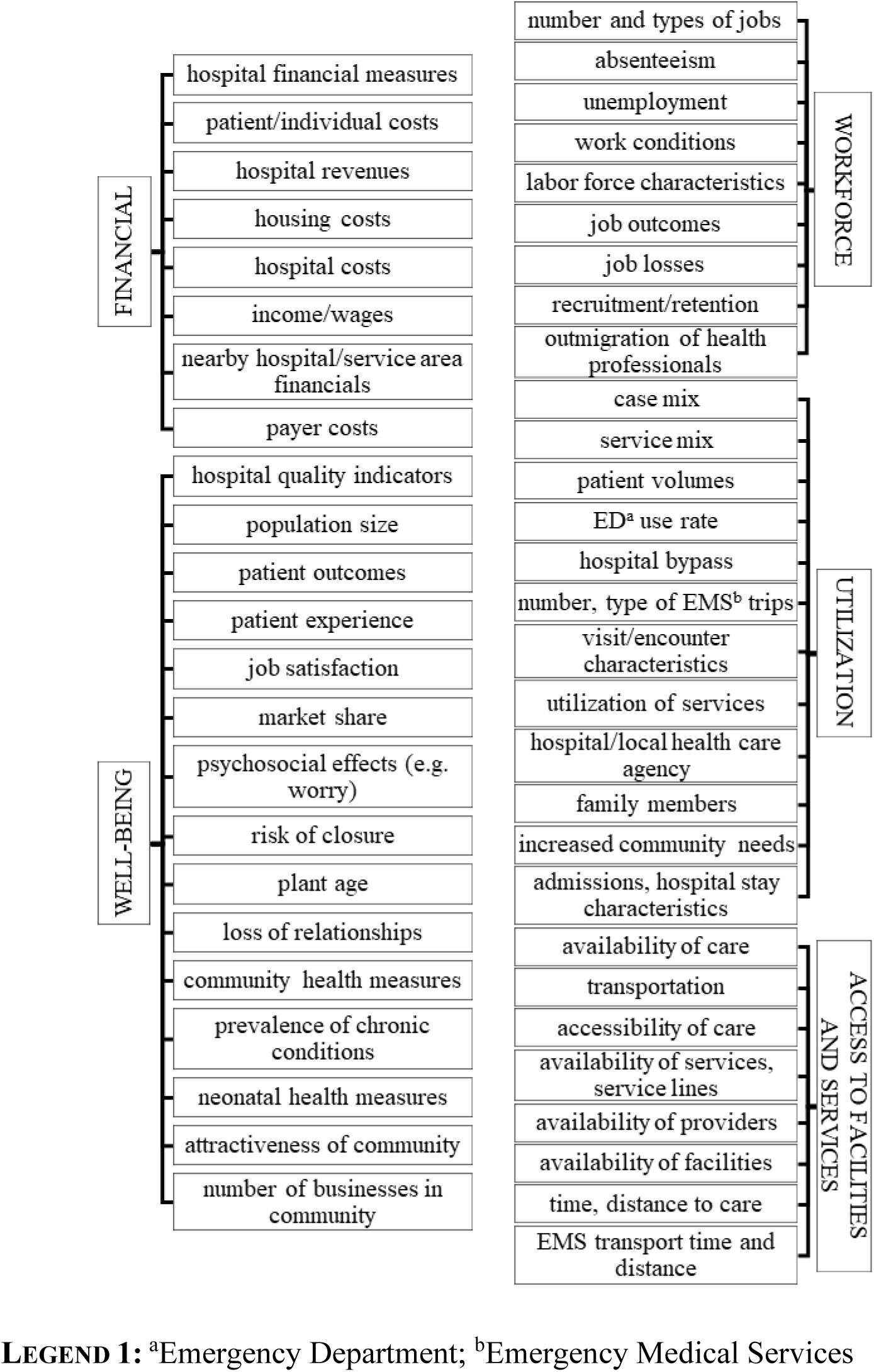
Outcome measures reported in included studies, grouped into categories.

In the second step, we categorized the affected entities that experienced studied outcomes. These were varied and complex, but overall fell into two distinct categories: persons and their communities, and health care institutions and their environments (Figure 2).

**Figure 2:**
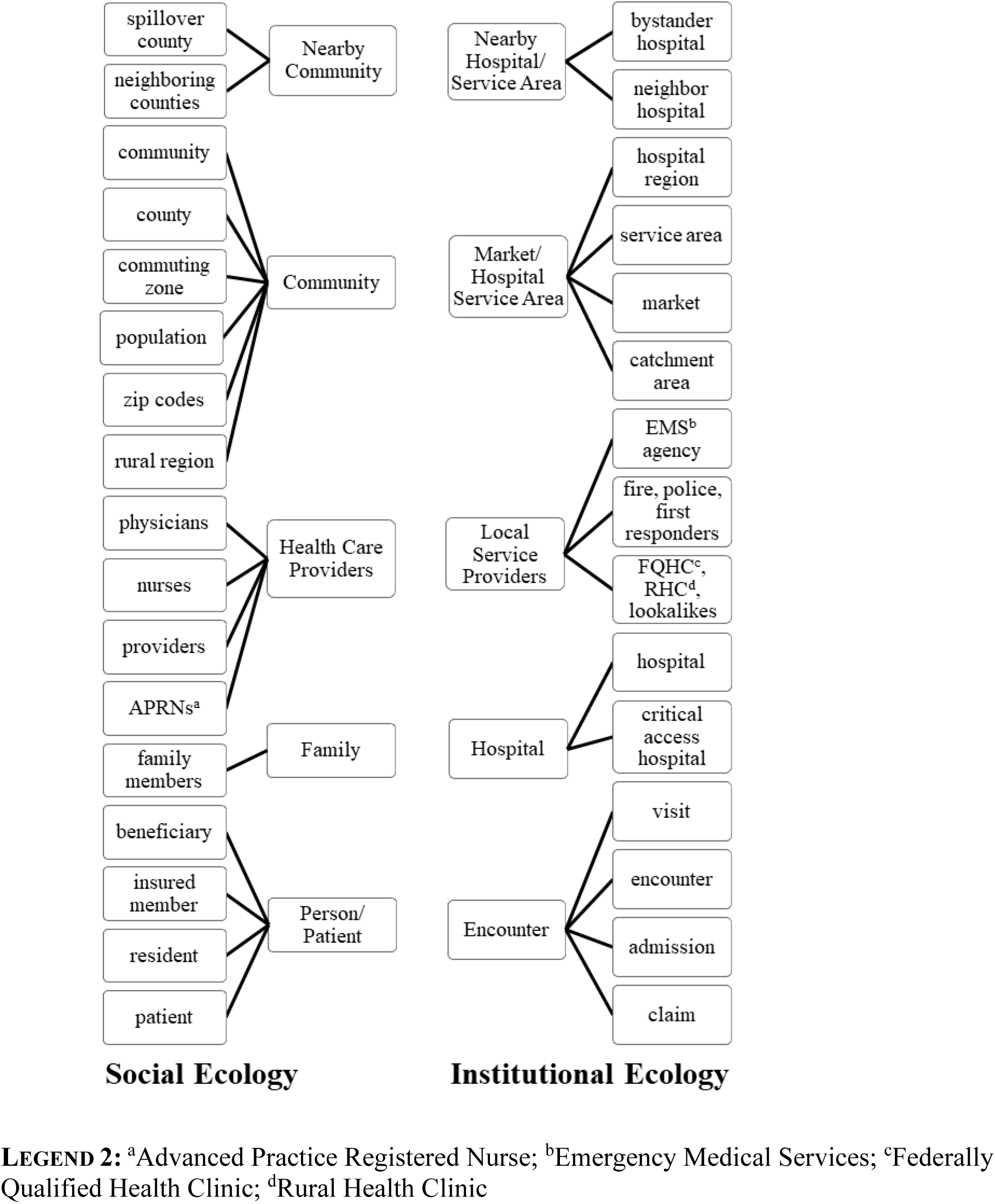
People, groups, or entities experiencing outcomes of rural hospital closures or mergers, coded into inductively identified groups.

In our third and final step, we combined our categories of outcomes and affected entities into a matrix which is depicted and described in the results section of this paper. We named the framework the *Health System Ecologies Impact Matrix*.

#### Phase 2: Applying the framework as a tool for epistemic critique

We wished to explore what we do and do not know about the impact of rural hospital closures and mergers to understand whose knowledge and experience is valued and legitimized through research. To do this, we systematically classified the impacts (outcome-affected entity pairs) reported in each included article and tabulated them using our newly derived impact matrix. To ensure that the coding was rigorously applied in the analysis, three of the researchers (AC, JP, KS) coded the outcomes and the affected entities for a subset of five articles. We discussed discrepant results, refined descriptions of codes, and repeated the coding process with a second set of five articles to ensure agreement. A single researcher (AC) coded the remainder of the dataset, randomly selecting approximately 20 percent of the studies to also be coded by the other two coders (JP, KS). Each outcome reported in a paper was mapped onto the impact matrix which we used to guide our analysis of what has been studied within this literature and to identify knowledge gaps where no outcomes have been reported.

## Results

We first describe the inductively derived framework (phase 1) and then we summarize what we do and do not know about rural hospital mergers and closures (phase 2). Throughout, we use the term “outcome” or “outcome measure” to indicate what is measured and reported by researchers in relation to closure or merger, “affected entity” to identify who or what experiences the outcome, and “impact” to denote related sets of outcomes and affected entities. A detailed description of the results of the scoping review search and selection is available in the Supplementary Material. Figure 3 depicts a flowchart of the search and selection process. Characteristics of the 46 included primary studies are summarized in Table 1.

**Figure 3:**
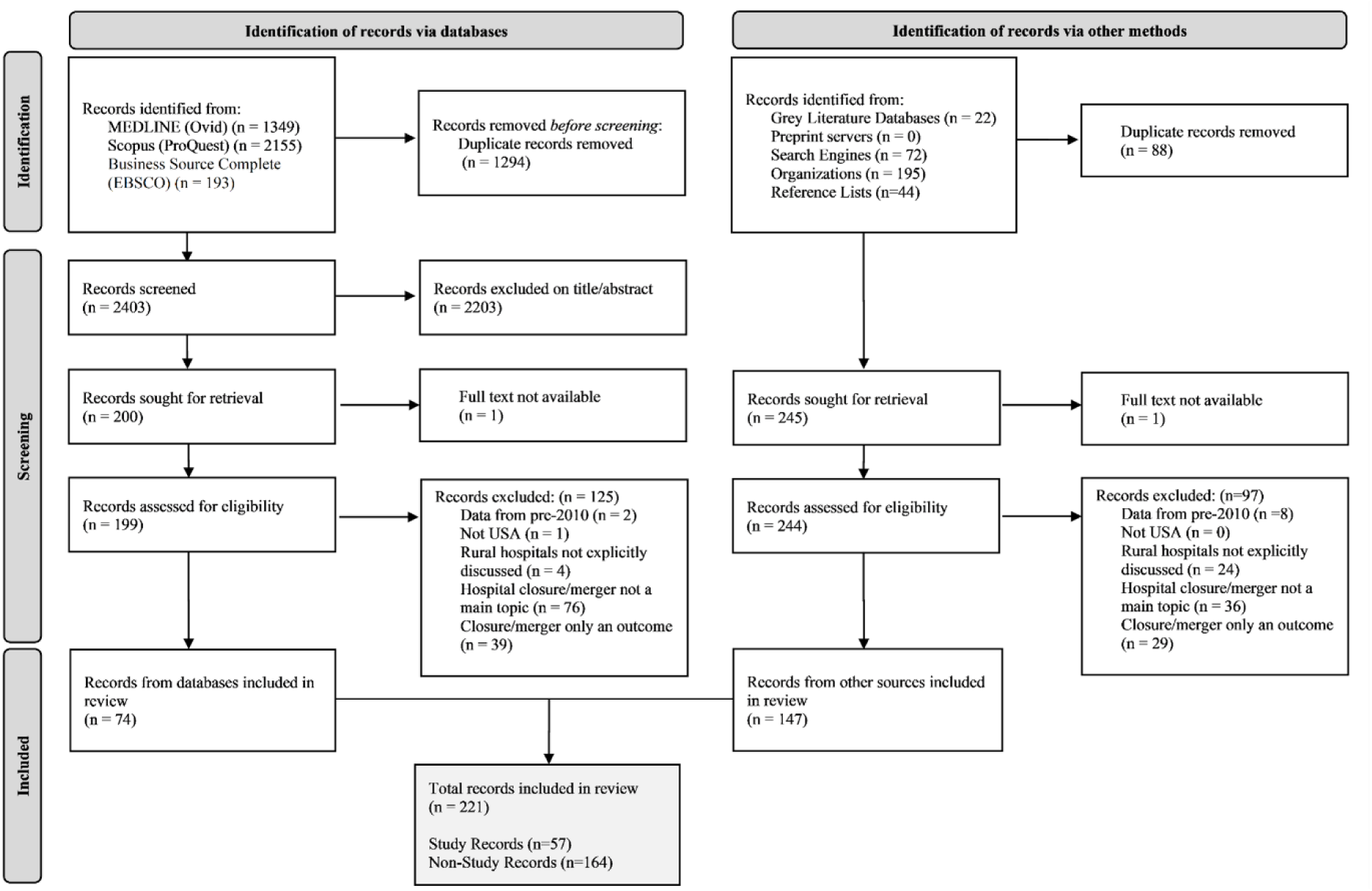
PRISMA flow diagram illustrating record selection process.

**Table 1:** Selected characteristics of included primary studies.

|  | *Total* | *Studies Related to Closures* | *Studies Related to Mergers* |
| --- | --- | --- | --- |
| **Methods** |  |  |  |
| *Quantitative* | 37 | 27 | 10 |
| *Qualitative* | 6 | 6 | 0 |
| *Mixed or multiple methods* | 2 | 2 | 0 |
| *Other* | 1 | 1 | 0 |
| **Type of Publication<sup>a</sup>** |  |  |  |
| *Journal article* | 26 | 18 | 8 |
| *Dissertation* | 6 | 5 | 1 |
| *Non-journal report* | 23 | 20 | 3 |
| *briefs* | 7 | 4 | 3 |
| *reports* | 14 | 14 | 0 |
| *other* | 2 | 2 | 0 |
| **Geographies** |  |  |  |
| *National* | 31 | 23 | 8 |
| *Other (e.g. state, county, health system, etc.)* | 15 | 13 | 2 |
| **Sources of Data<sup>b</sup>** |  |  |  |
| *Administrative data* | 38 | 29 | 9 |
| *Primary data* | 10 | 9 | 1 |

<sup>a</sup>Some studies yielded multiple publications so numbers may not total 46
<sup>b</sup>Some studies used multiple data sources so numbers may not total 46

### Phase 1: A novel framework for understanding health system impacts

We developed an analytical framework, grounded in the rural hospital merger and closure data, for understanding the impacts of health system changes.

In our first phase of analysis, we identified five categories of outcomes: financial, workforce, utilization, well-being, and access to facilities and services. We identified the people, groups, or entities that were reported to experience a given outcome: patients and individuals, families, health care providers, communities, neighboring communities, encounters or admissions, hospitals, non-hospital service providers, hospital or health care markets or service areas, and neighboring hospitals or service areas. We observed a substantial variety in outcomes captured within each category. While some categories of impact are relatively similar across levels (e.g. what constitutes a “financial” impact), others manifest differently for different affected entities (e.g. what constitutes a “well-being” impact).

We found that sources generally examined outcomes of hospital mergers and closures on affected entities from one of two perspectives. One focused on how hospital closures impact individual people, patients, their families and caregivers, health care workers, the communities they live in, and the communities that neighbor them – the *social ecology*. The other set examined encounters and visits, hospitals, health service organizations, hospital markets and service areas, and nearby hospitals or service areas, which we considered to be the *institutional ecology*. Constructing two separate but somewhat parallel ecologies brought clarity to our analysis: rather than an eclectically assembled matrix of outcomes too diverse to synthesize, we discovered coherence within each ecology.

When discerning between the impacts reported within the two ecologies, some outcomes appear similar but differ in important ways. For example, five studies (Gujral & Basu, 2019; Heard et al., 2022; Jiang et al., 2021, 2022; Khushalani et al., 2022) all looked at length of stay and mortality as outcomes of utilization and quality, respectively, but they studied them in different ways. In research on rural hospital closures, Khushalani (2022) looked at these outcomes within the service area of the closed hospital (institutional) while Gujral & Basu (2019) studied privately insured patients within a given geographic area (social). In examining impacts of hospital mergers, Jiang et al. looked at length of stay and in-hospital deaths at the hospital level (Jiang et al., 2021, 2022) while Heard et al. (2022) studied the same at the encounter level (all within the institutional ecology).

Some researchers examined affected entities which appeared similar despite being classified into different ecologies. For example, several studies looked at geographic regions of some kind. On the social side, especially for quantitative studies, researchers tended to study counties – administrative units associated with the places where people live. Qualitative studies, more tolerant of fuzzy boundaries for their regions or communities, still typically related to a social grouping of people bound, at least in this set of literature, by common geography. On the institutional side, similar geographic regions were often delineated relative to hospitals (e.g., rationally constructed hospital service areas or catchment areas, or logically defined markets). Our Health System Ecologies Impact Matrix is depicted in Figure 4.

**Figure 4:**
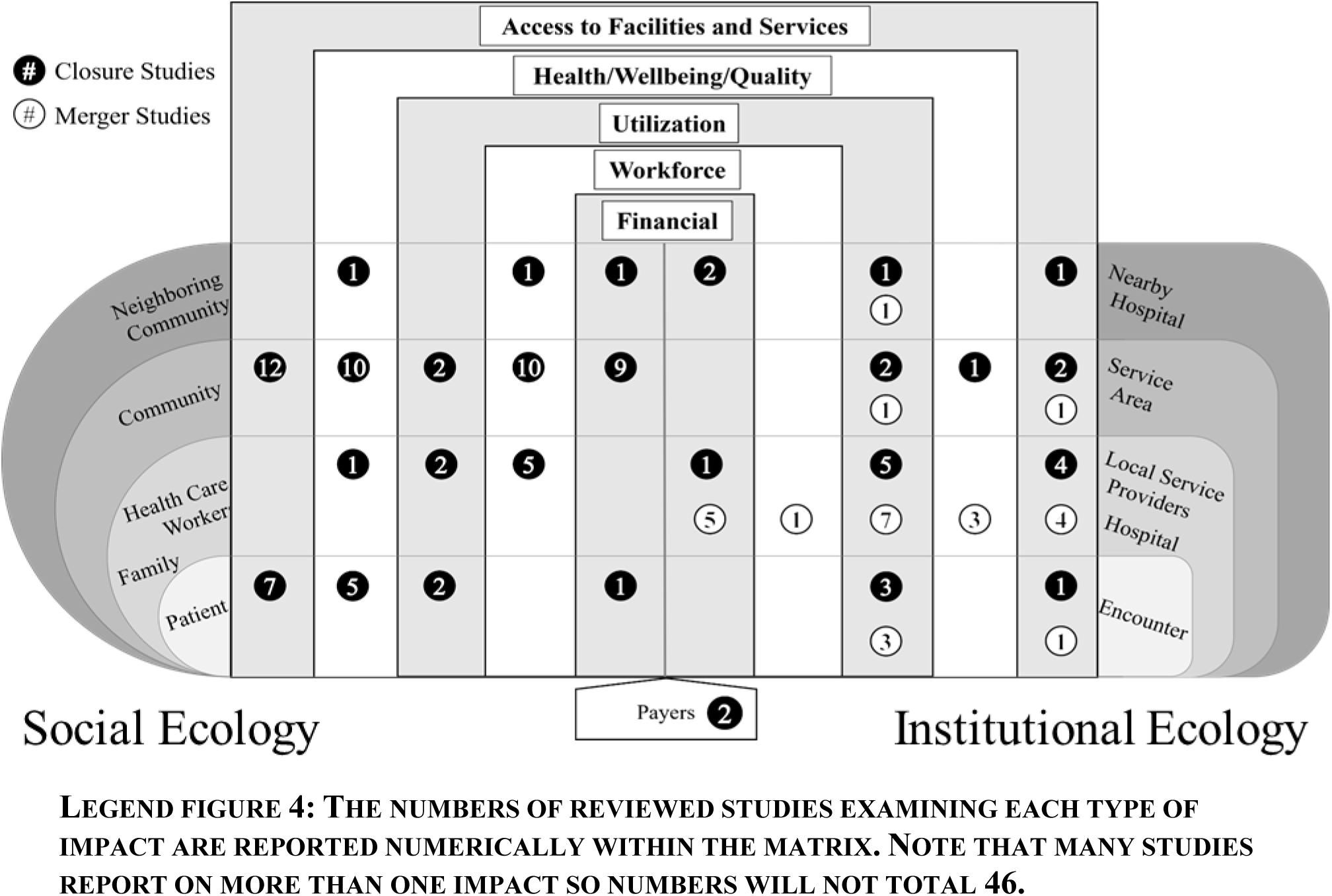
Health System Ecologies Impact Matrix.

### Phase 2: Application of the Impact Matrix to the included literature

In Figure 4, we indicate the number of studies (closure studies in black circles and merger studies in white circles) that explored outcomes within each of the impact categories for the intersecting affected entity. (In the supplementary material we provide two variations of this figure – one for hospital closures and one for mergers – with study references listed in lieu of counts).

In the following two sections, we describe the literature that reports on studies of rural hospital closures and on studies of rural hospital mergers. For each set of literature (closures and mergers), we first provide descriptive characteristics of the included studies and reflect on the overall state of the literature. We then narratively summarize what we know (the impacts reported in the literature) and what we do not know (areas within the matrix with no reported impacts) organized by category of outcome.

#### Effects of rural hospital closures

Of the 46 studies included in this review, 36 examined outcomes related to rural hospital closures. A table capturing the full list of included studies evaluating impacts of rural hospital closures is found in the Supplemental Material; conclusions are reported for qualitative studies and for quantitative studies where results showed statistical significance.

Most of the included closure studies (27/36, 75.0%) employed quantitative approaches using state or national administrative datasets to explore impacts. Qualitative methods were the most common approach in studies that examined patient or person-level impacts (Letheren et al., 2024; Medicare Payment Advisory Commission (MEDPAC), 2021; Mike, 2020; J. G. Smith et al., 2022; Wishner et al., 2016). Qualitative studies, or the qualitative components of mixed or multiple method studies, also produced findings related to communities (Capital Link, 2022; Medicare Payment Advisory Commission (MEDPAC), 2021; Tennessee Health Care Campaign, 2021; Wishner et al., 2016), health care workers (Letheren et al., 2024; J. G. Smith et al., 2022; Thomas et al., 2015; Wishner et al., 2016), family members (Mike, 2020), or regarding local non-hospital service providers (Medicare Payment Advisory Commission (MEDPAC), 2021; Tennessee Health Care Campaign, 2021; Thomas et al., 2015). These qualitative studies relate predominantly to the social ecology.

The lack of evaluation of hospital-level outcomes is unsurprising since in the case of closure, no hospital exists to experience effects. Very few studies looked at the effects of hospital closures on surrounding communities (Alexander & Richards, 2023). nearby hospitals (Ramedani et al., 2022; L. D. Song & Saghafian, 2019), or neighboring service areas (L. D. Song & Saghafian, 2019).

##### Financial

Financial outcomes were reported in 13 of the 36 closure studies (36.1%) and were most examined at the community level (9/13, 69.2%). Of the studies that reported outcomes relevant to the community, most reported county-level economic data related to income or wages (Chatterjee, 2022; Edmiston, 2019; Eilrich et al., 2015; T. L. Malone et al., 2022; Manlove & Whitacre, 2017; Tennessee Health Care Campaign, 2021; Thomas et al., 2015; Vogler, 2020), consumer financial health (Alexander & Richards, 2023; Chatterjee, 2022), and housing-related costs (Alexander & Richards, 2023; Manlove & Whitacre, 2017; Vogler, 2020). One paper examined the effects of hospital closure on consumer financial health and housing costs for communities experiencing closure as well as for their neighboring communities (Alexander & Richards, 2023). Only one study reported on financial impacts to individual patients following hospital closure (Letheren et al., 2024) – namely, lack of predictability of costs and affordability of care. Two papers investigated spillover effects of hospital closures on neighboring hospital or service area financial measures (e.g. profitability, costs of care, and operational efficiency) (Ramedani et al., 2022; D. (Lina) Song, 2020). A single study reported increased operational costs for local Emergency Medical Services (EMS) providers, which aligns with the non-hospital service provider group (Eilrich et al., 2015). Two studies examined increased spending for payers (i.e., private insurance (Andreyeva et al., 2022) and Medicare (US Government Accountability Office, 2020)) following hospital closure.

Within the dataset, no studies examined financial impacts of hospital closures for families and health care workers, or for the hospital, encounter, or health service area for the hospital that closed.

##### Workforce

Workforce-related outcomes were examined by 13 of the 36 included studies (36.1%) and were again most commonly examined at the community level (9/13, 69.2%). Community workforce outcomes (e.g. data about jobs, employment, unemployment, occupational and industry mix, etc.) were often examined in tandem with aforementioned quantitative county-level financial and economic analyses (Alexander & Richards, 2023; Chatterjee, 2022; Edmiston, 2019; Eilrich et al., 2015; T. L. Malone et al., 2022; Manlove & Whitacre, 2017; Vogler, 2020). Qualitative research provides rich accounts related to job losses and related increases in unemployment (Thomas et al., 2015), reported outmigration of health professionals (Wishner et al., 2016), and discussions of the impact of hospital closure on non-healthcare jobs within the community (Wishner et al., 2016). One study examined employment-related impacts on neighboring communities (Alexander & Richards, 2023). Five studies looked specifically at health worker related employment impacts, evaluating changes in numbers of health care jobs (Alexander & Richards, 2023) or the supply of health care providers (Letheren et al., 2024; Mobley et al., 2020; Wishner et al., 2016), and providing qualitative contributions about the impacts on nursing jobs and job outcomes (J. G. Smith et al., 2022).

No studies reported workforce-related impacts at the patient level, nor were workforce impacts investigated at any level within the institutional ecology.

##### Utilization

Utilization of facilities and services was examined in 16 (44.4%) of the 36 included closure studies. Within the institutional ecology, utilization has been studied at all levels. Three studies looked at the encounter level, two of which examined changes in EMS utilization in terms of number of trips and proportion of non-emergent trips (Nikpay et al., 2021) or in terms of ambulance call times (Chaudhary et al., 2018). The other examined commercial insurance claims to evaluate changes in number of health care visits of various types: outpatient, inpatient, ED without admission, ED with admission, and visits with primary care or specialist providers (Andreyeva et al., 2022). Other studies looked at the impacts of hospital closures on EMS at the service provider level, reporting on aggregated or average EMS transportation time or mileage (Eilrich et al., 2015; Nikpay et al., 2021) and average EMS response times (Medicare Payment Advisory Commission (MEDPAC), 2021). Quantitative findings from these studies are complemented by qualitative studies that highlight increased stress or burden on EMS (Tennessee Health Care Campaign, 2021; Thomas et al., 2015) and other emergency service providers (Tennessee Health Care Campaign, 2021).

Two studies looked at the health service areas of closed hospitals to look at residents’ utilization of health services. One study examined admission rates and average length of stay for emergency care sensitive conditions (ECSC) and ambulatory care sensitive conditions (ACSC) (Khushalani et al., 2022). Another looked at the number of inpatient admissions, hospital outpatient visits, and the number of evaluation and management (E&M) visits across settings (Medicare Payment Advisory Commission (MEDPAC), 2021). A single study examined utilization impacts to bystander hospitals in terms of hospital admissions and ED visits (Ramedani et al., 2022).

At the person or patient level of the social ecology, one study reported on birthing women’s rates of cesarian delivery, induction and in-hospital births (Durrance et al., 2024) and another looked at length of stay within a population of insured patients (Gujral, 2020). One qualitative study found increased utilization of family members for the provision of health-related tasks including transportation to access care (Mike, 2020) and another reported an increased workload or increased utilization of the healthcare providers who remained in the community after hospital closure (Letheren et al., 2024). Two studies looked at county-level utilization outcomes among Medicare beneficiaries: numbers of emergency department (ED) visits (Gelbaugh & Advisory Board, 2021; US Government Accountability Office, 2020), inpatient stays (US Government Accountability Office, 2020), and outpatient visits (US Government Accountability Office, 2020).

No studies examined impacts of hospital closures on neighboring communities after a rural hospital closed.

##### Access to Facilities and Services

Within the social ecology, most of the studies exploring changes in access to services and facilities following hospital closure focused on patients or communities. One study took a geospatial approach to determine the number and proportion of people who can access a hospital within specific drive-time intervals (McCarthy et al., 2021). Patient access to care was often discussed in terms of time or distance needed to access care (Letheren et al., 2024; Medicare Payment Advisory Commission (MEDPAC), 2021; Mike, 2020; Wishner et al., 2016), sometimes accompanied by a description of transportation-related challenges (Letheren et al., 2024; Wishner et al., 2016), increased wait times (Letheren et al., 2024), or patients delaying or forgoing needed care (Thomas et al., 2015; Wishner et al., 2016). Some studies reported on access to care broadly (Mike, 2020; J. G. Smith et al., 2022; Wishner et al., 2016) or with respect to specific health care services, such as emergency care (Letheren et al., 2024; Wishner et al., 2016) or specialist care (Letheren et al., 2024; Tennessee Health Care Campaign, 2021; Wishner et al., 2016). One study looked at the likelihood of a person giving birth in their county of residence (Durrance et al., 2024) as indicative of their access to labor and delivery services post hospital closure.

Twelve (33%) of the closure studies looked at access to facilities and services at the community level. Surprisingly, only one study examined the impacts of closures on a region’s access to a hospital (i.e., distance or time to the nearest hospital) (US Government Accountability Office, 2020). Four studies reported on access to health care providers within a community, looking at physicians, generally and by specialty (Germack et al., 2019; Mobley et al., 2020; Tennessee Health Care Campaign, 2021; US Government Accountability Office, 2020), and at Advanced Practice Providers (APPs) or Advance Practice Registered Nurses (APRNs) (Germack et al., 2021; Mobley et al., 2020; US Government Accountability Office, 2020). Several studies explored access to non-hospital facilities and services like community health services (i.e., Federally Qualified Health Centers (FQHC), Community Health Centers (CHC) and “lookalike” organizations) (Bell et al., 2023; Capital Link, 2022; Medicare Payment Advisory Commission (MEDPAC), 2021; Miller et al., 2021; US Government Accountability Office, 2020), pharmacies (Capital Link, 2022), freestanding EDs (Medicare Payment Advisory Commission (MEDPAC), 2021), nursing homes (Mills, 2022), and urgent care facilities (Medicare Payment Advisory Commission (MEDPAC), 2021).

Relatively few studies explored access to care within the institutional ecology. A few studies about ambulance and EMS reflect changes in access *to* and *by* these services. At the encounter level, access *to* EMS services is reflected in ambulance response times (Miller et al., 2020). Changes in average or total transport times and distances (Chaudhary et al., 2018; Medicare Payment Advisory Commission (MEDPAC), 2021; Nikpay et al., 2021; T. B. Smith et al., 2022) illustrate differences in access *to* hospitals *by* the EMS providers. Two papers looked at the hospital service areas of closed hospitals to understand impacts on access to various hospital-based services (e.g., mental health services, substance use disorder treatments, cancer-related services, etc.) (US Government Accountability Office, 2020; Zahnd et al., 2023).

No outcomes related to access to care were reported at the level of families and health care workers, or neighboring communities, nor were any reported relative to neighboring hospital markets.

##### Well-being

Outcomes of hospital closures which reflect well-being are diverse, often aligned with measures of health or quality. Outcomes in this category have been most commonly studied in the social ecology, manifesting along a variety of dimensions: the strength of the housing market (also studied for the neighboring counties) (Alexander & Richards, 2023), stability or growth in the population (Chatterjee, 2022; T. L. Malone et al., 2022), levels of job creation (Tennessee Health Care Campaign, 2021), community prestige or morale (Capital Link, 2022; Eilrich et al., 2015; J. G. Smith et al., 2022), or in a community’s ability to attract new residents (Tennessee Health Care Campaign, 2021; Vogler, 2020; Wishner et al., 2016) or new businesses (Vogler, 2020; Wishner et al., 2016). Of course, community well-being can also be reflected in measures more closely aligned with health, such as disability program participation (Chatterjee, 2022) or county-level inpatient and 30-day mortality rates (Merrell, 2019) (the latter two being more commonly used as hospital quality measures rather than community-level measures).

At the patient level, many of the studies looked at biomedical health measures: neonatal health outcomes (i.e. low birthweight, number of preterm births, Apgar scores, and infant mortality) (Durrance et al., 2024), prevalence of chronic conditions (US Government Accountability Office, 2020), or inpatient mortality (Gujral & Basu, 2019). One qualitative study revealed a loss of familiar connections with health care providers that resulted in a poorer experience of care (Letheren et al., 2024) and another discussed the psychosocial effects felt by individuals affected by hospital closure (J. G. Smith et al., 2022). Only one study reported on the effects of hospital closures on the well-being of health care workers – namely, the psychosocial effects on nurses (J. G. Smith et al., 2022).

Few studies examined well-being impacts within the institutional ecology. One study looked at in-hospital mortality within the closed hospital’s service area (Khushalani et al., 2022), while another examined 30-day mortality and 30-day readmission at nearby hospitals (Nikpay et al., 2021).

No studies examined well-being related impacts of hospital closures at the local service provider level, or at the encounter level.

#### Effects of rural hospital mergers

Of the 46 studies included in this review, ten (21.7%) examined outcomes related to rural hospital mergers, all of which were framed within the institutional ecology. Every study on the topic of hospital mergers employed quantitative methods, usually using national administrative datasets. Almost all of the included research on hospital mergers looked at impacts on the merged rural hospital. Surprisingly, no studies looked at the impacts of rural hospital mergers on the acquiring hospital or parent health network. A table detailing the full list of included studies evaluating the impacts of rural hospital mergers is found in the Supplemental Material.

##### Financial

Five of the ten studies (50.0%) about hospital mergers reported on financial outcomes related to merged hospitals. Traditional financial performance measures form the basis of several studies (e.g., costs or expenses (Jiang et al., 2022; Noles et al., 2015; Williams Jr. et al., 2020), revenues (Noles et al., 2015; D. J. Williams et al., 2021; Williams Jr. et al., 2020), and profitability ratios and margins (Noles et al., 2015; O’Hanlon et al., 2019; Williams Jr. et al., 2020)). Two studies look specifically at capital expenditures post-merger (Noles et al., 2015; Williams Jr. et al., 2020). Two studies look at hospital financial health or strength through rates of hospital financial distress (Jiang et al., 2022), market share (Jiang et al., 2022), and ability to cover debt (Williams Jr. et al., 2020)).

No studies examined any impacts at any other level within the institutional ecology.

##### Workforce

Only one of the ten studies (10.0%) reported on changes in workforce-related factors after hospital merger: the number of full-time equivalents (FTEs) per bed, and the average salary of an FTE (Noles et al., 2015).

No workforce-related outcomes were reported at the encounter level, nor for merged hospitals’ service area, nor for nearby hospitals.

##### Utilization

Nine of the ten included merger studies (90.0%) examined at least one outcome related to utilization. Two studies look at the encounter level. One reported on the number of hospital stays within certain categories (Jiang et al., 2021); the other, a single-institution study, looked at changes in trauma service utilization (e.g., injury severity, number of days in hospital, in intensive care, or on a ventilator, and number of specialist consults) (Heard et al., 2022). Seven studies examined hospital utilization post-merger as measured by number of admissions, inpatient stays or discharges (Henke et al., 2021; Jiang et al., 2022; Noles et al., 2015; O’Hanlon et al., 2019; D. J. Williams et al., 2021; Williams Jr. et al., 2020), length of stay (Jiang et al., 2022), number of beds (Jiang et al., 2022), average daily census (Noles et al., 2015; D. J. Williams et al., 2021; Williams Jr. et al., 2020), and service mix (Jiang et al., 2022). One study looked at inpatient admissions within certain diagnostic categories to understand the proportion of stays that bypass the local hospital (T. Malone et al., 2020) and another evaluated non-inpatient hospital utilization for ED and non-emergency purposes (O’Hanlon et al., 2019).

##### Access to Facilities and Services

Four studies focused on access to facilities and services following the merger of a rural hospital with a health system. Most commonly, researchers reported on the availability of specific technologies (O’Hanlon et al., 2019) or service lines (Henke et al., 2021; O’Hanlon et al., 2019; Oyeka et al., 2023). In one study, researchers examined the distance travelled by patients admitted to the hospital (Jiang et al., 2022).

No studies examined access at the level of the encounter, the service area, or the neighboring hospital.

##### Well-being

Five studies report on well-being-related measures post rural hospital merger.

Hospital well-being was evaluated by one research team using average age of assets, with “healthier” hospitals falling within the newest quartile (Williams Jr. et al., 2020). Another researcher considered hospital well-being as manifesting through its risk of closure (Jiang et al., 2022). One paper reported on hospital quality as a function of patient experience and 30-day readmission rate (O’Hanlon et al., 2019). Mortality-related metrics were reported at the encounter level (Heard et al., 2022; Jiang et al., 2021). Also at the encounter level, one study reported on elective procedure complication rates (Jiang et al., 2021) and another examined if there were differences in proportions of patient disposition following trauma admission (Heard et al., 2022).

No studies examined well-being impacts for the merged hospital’s service area or for neighboring hospitals.

## Discussion

The paucity of journal articles and prevalence of non-academic publications, especially related to the long-standing problem of rural hospital closures, illustrates some of the challenges that rural researchers face in disseminating their work. More than half of the studies included in our review were published outside of the peer reviewed literature. One potential reason for this imbalance might be that some academic journals with national or international audiences prefer not to publish research that is too locally situated, whose relevance is not seen as adequately applicable to the health system at large (Hyland, 2015). For many rural health systems stakeholders, differentiation from the urban-dominant US health system is the point: what knowledge holds true at the national level may not among individual rural communities across the country. Operating within a disciplined approach to research and publication constrains the types of people who may contribute, the types of evidence that are considered legitimate, and the types of findings that are given weight (Hyland, 2015; Hyland et al., 2023). Restricting a search to only academic sources shapes a world view that restricts the interpretation of reality through only a privileged (and often urban-centered) lens.

Had we restricted our search to only peer reviewed literature, our understanding of rural hospital closures, especially, would have been narrower. For many rural researchers, academic and funding priorities incentivize rapid reporting of sponsored reports, often in non-academic outlets. Mullens et al. (2024), who excluded gray literature from their recent scoping review on rural hospital closures, noted a lack of evidence about changes in access to specialty care which we found discussed in several studies (Mobley et al., 2020; Tennessee Health Care Campaign, 2021; US Government Accountability Office, 2020; Wishner et al., 2016). Gray literature offered our only insights into the impacts of closures on community-level utilization, EMS agency financial metrics, and quality measures of bystander hospitals.

Disciplinary conventions that dictate what constitutes appropriate evidence (Hyland et al., 2023) may explain the dominance of quantitative methods and the prevalence of use of administrative data sources within the included literature, especially from academic journals. Data found in publicly accessible government datasets convey a pre-vetted reliability that lends weight to the findings they generate. Administrative data sets are not inherently unbiased or politically neutral; what gets measured, what categories are allowed, and the granularity or scale at which data are aggregated are all determined by the systems that construct and fund the data collection. Problems with the collection of gender and racial data continue to trouble health researchers (Hardeman & Karbeah, 2020; Johnson et al., 2023; Richards et al., 2022), though considerable efforts are underway to develop antiracist approaches to administrative dataset research (Follis et al., 2023; Hardeman et al., 2022; Krieger, 2021; McLemore, 2021).

Administrative data are collected to fit a purpose. Claims data from a private healthcare payer, for example, will capture data that are deemed important by health insurers and their boards of directors, regardless of whether these align with researcher or public interest. Datasets that report on services delivered (e.g., Medicaid and Medicare utilization reports or electronic health record data) are biased toward people who have access to care; people who delay or forego care, as noted in some of the qualitative research we reviewed, are invisible in such analyses. Health services utilization data should also not be used as a proxy for understanding health care needs. Utilization reflects needs only in a situation of perfect health equity – where people who need care can access it, physically, financially, and safely. Using utilization data to characterize the health needs of a community where access is restricted (e.g., because there are not enough providers, no nearby services, no insurance coverage, no transportation, etc.) is problematic, and we know rural communities fall into several of these categories. The dominant modes rural researchers use to study the impacts of hospital closures and mergers – namely, quantitative studies using large administrative datasets – shape the types of knowledge that emerge from this research.

### What we can learn about hospital closures and mergers through the lens of social and institutional health system ecologies

Most of the studies that examined outcomes of rural hospital closures were explored within the social ecology and community-level impacts which dominated that research. That so many of the quantitative studies on hospital closures reported on communities likely reflects the ready availability of county-level data. The implicitly social nature of communities explains why several qualitative studies also embrace this unit of analysis. Evidence about how hospital closures affect communities addressed all five categories of impacts with access to care being most extensively studied.

Qualitative studies reported almost exclusively within the social ecology. Researchers using qualitative methods captured voices not otherwise represented in the corpus of hospital closure studies: the concerns of nurses whose hospitals closed (J. G. Smith et al., 2022), reflections from patients about the extra burden shouldered by their families (Mike, 2020), and patient perceptions about changing costs of care (Letheren et al., 2024). Qualitative work revealed findings that are unmeasured (or unmeasurable) in quantitative administrative data, such as psychosocial impacts on individuals (Capital Link, 2022; J. G. Smith et al., 2022; Tennessee Health Care Campaign, 2021), health care providers, and communities (Capital Link, 2022; Eilrich et al., 2015; Thomas et al., 2015), increased wait times to access care (Letheren et al., 2024), delayed or foregone care (Thomas et al., 2015; Wishner et al., 2016), or impacts on interpersonal relationships (Letheren et al., 2024).

No studies of rural hospital mergers reported outcomes within the social ecology. Given that the research on mergers has all been published within the last five years, neglect of person-centered outcomes may reflect a nascent field. The dominance of peer reviewed journal articles within this subset of literature, however, may suggest a disciplinary bias towards thinking about mergers as a purely rational, institution-oriented phenomenon. Future research on how rural hospital mergers with larger health systems affect patients, people, health workers, and communities is critical given the steep rise in affiliations in recent years.

Within the institutional ecology, utilization has been the most researched impact domain across both closure and merger studies. Data availability (i.e., the accessibility of claims data, hospital electronic health records, and the trend towards centralized data reporting for EMS providers) helps to explain the prevalence of utilization outcomes. Other than one merger study which reported on the number of FTEs per bed and salary expenses – an illustrative example of an institution-oriented view of the role of humans within a system – no workforce issues were reported for institution-centered stakeholder groups. Despite being the most studied impact domain on the person-centered side, relatively few studies explored access to care within the institution-centered discourse.

### How to know what we do not know: a tool for identifying knowledge gaps

The Health System Ecologies Impact Matrix visually depicts knowledge gaps, or in other words, shows us what we do not yet know. Using the matrix as a visual research tool permits us to compare the completeness of the knowledge base within or across topics, illuminating gaps or insufficiencies in current knowledge. For example, inspecting the matrix of impacts of rural hospital closures shows that workforce issues related to hospital closure are completely un-studied within the institutional ecology. While some vacancies in the matrix are self-explanatory (e.g., we would not expect hospital closures to have workforce effects at the encounter level), others reveal opportunities for future research (e.g., workforce-related outcomes should be explored within a health service area or at a nearby hospital following rural hospital merger or closure).

We believe that classifying literature into the matrix permits us to appraise the completeness of our knowledge and to identify areas where research is still needed. This tool may be beneficial to researchers, health care managers, and policymakers as they embark on new research endeavors, consider changes to their organizations, or anticipate effects of potential policy changes related to the health care delivery system.

### Limitations and future research

We developed the Health System Ecologies Matrix based on literature about rural hospital closures and mergers; however, none of the elements of the framework specifically relates to rural contexts, to hospitals, nor to types of health system changes. We believe that this tool – or adaptations thereof – may be more widely applicable to other types of health system changes (e.g., new facility openings, facility relocations, structural changes, etc.) and may be applied relative to other types of health organizations (e.g., nursing homes, primary care offices, etc.). Future research could use the Health System Ecologies Impact Matrix to assess a different body of literature.

Equity considerations are not explicitly captured in our framework. Few outcome measures – at least from our current dataset – truly measure degree or quality of equity. More commonly, studies report on ways that outcomes may be differently felt by vulnerable people or racialized groups, for example. Many of our included articles accounted for equity-related factors in some way, whether through sensitivity or subgroup analyses aligned with social stratifying factors known to impact health outcomes (O’Neill et al., 2014) or through research questions aligned to health equity. We hope that future applications of the Health System Ecologies Impact Matrix in larger bodies of literature might suggest an appropriate way to incorporate equity into the model.

## Conclusions

Our novel framework for understanding what we do and do not know about the impacts of health system changes is, to our knowledge, the first that permits the classification of outcomes by category of outcome and by affected entity. Applying the Health System Ecologies Impact Matrix to systematically reviewed literature permits a rapid appraisal of the extent and character of existing knowledge and easy identification of opportunities for additional research. Such models may also help policymakers seeking to make informed decisions or plan future research funding initiatives.

## Acknowledgements

We acknowledge the assistance of Emily Da Silva, MIS, University of Ottawa Library, for feedback and guidance in developing the search strategy. We are also grateful to Mary Cabral, Health Sciences Librarian at Clarkson University, for her collaboration in providing a peer review of the electronic search strategy.

## Statements and Declarations

### Ethical considerations

not applicable

### Consent to participate

not applicable

### Consent for publication

not applicable

### Data availability

The data underlying this review are included or referenced in this published article and its supplementary information files. Datasets generated from the underlying data which support our analysis are available from the corresponding author on reasonable request.

