## Supplemental Material for "The Impact of Rural Hospital Closures and Mergers on Health System Ecologies: A Scoping Review"

### Online Supplemental Material

### Scoping Review Methods

#### Search Methods

In consultation with an academic librarian, we developed a comprehensive search strategy to incorporate appropriate key terms, synonyms, and related subject headings pertaining to two main concepts: *rural hospitals* and *closures or mergers*. The search was developed and piloted in Medline and was peer reviewed(McGowan et al., 2016) before it was adapted for the other included databases, Business Source Complete (EBSCO) and Scopus (ProQuest). The search was limited to articles in English published between January 2010 and December 2023. (Full search details are found in the Supplemental Material). Literature produced outside of the academic and commercial press – "gray literature" – was systematically searched with the assistance of a Grey Matters-inspired tool(CADTH, 2018) adapted to the US rural health context. Additional literature was identified by hand searching reference lists of included articles and through a focused search of Google and Google Scholar.(2020)

#### MEDLINE (OVID) Search Strategy

| # | **Query** |
|---|---|
| 1 | Rural Health/ or Rural Health Services/ or Rural Population/ |
| 2 | (rural* or ((remote or frontier) adj3 (area* or communit* or location*))).ti,ab,kf. |
| 3 | 1 or 2 |
| 4 | Hospitals/ or Hospitals, Low Volume/ or Hospitals, Community/ or Hospitals, General/ or Hospital Planning/ or Health Facilities/ or Financial Management, Hospital/ or Health Services Administration/ or Health Facility Administration/ or Hospital Administration/ |
| 5 | (hospitals or hospital or ((health or healthcare) adj3 (facility or facilities))).ti,ab,kf. |
| 6 | 4 or 5 |
| 7 | Hospitals, Rural/ or rural referral center.ti,ab,kf. |
| 8 | 3 and 6 |

| | |
|---|---|
| 9 | 8 or 7 |
| 10 | Health Facility Merger/ or Health Facility Closure/ or Multi-Institutional Systems/ or system affiliation.ti,ab,kf. |
| 11 | (merg* or consolidat* or closure* or close or closes or closed or closing or acquisition* or acquir*).ti,ab,kf. |
| 12 | 10 or 11 |
| 13 | 12 and 9 |
| 14 | limit 13 to (english language and yr="2010 -Current") |

#### Scopus

( ( TITLE-ABS-KEY ( merg* OR consolidat* OR closure* OR close OR closes OR closed OR closing OR acquisition* OR acquir* ) ) OR ( TITLE-ABS-KEY ( "system affiliation" ) ) ) AND ( ( TITLE-ABS-KEY ( "rural referral center" ) ) OR ( ( TITLE-ABS-KEY ( rural* OR ( ( remote OR frontier ) W/3 ( area* OR communit* OR location* ) ) ) ) AND ( TITLE-ABS-KEY ( hospitals OR hospital OR ( ( health OR healthcare ) W/3 ( facility OR facilities ) ) ) ) ) ) ) AND ( PUBYEAR > 2009 ) AND ( LIMIT-TO ( LANGUAGE , "English" ) )

#### Business Source Complete

Search Modes: Boolean/Phrase

| # | Query |
|---|---|
| S1 | TI rural* OR AB rural* OR KW rural* |
| S2 | TI ( (remote OR frontier) N3 (area* OR communit* OR location*) ) OR AB ( (remote OR frontier) N3 (area* OR communit* OR location*) ) OR KW ( (remote OR frontier) N3 (area* OR communit* OR location*) ) |
| S3 | S1 OR S2 |
| S4 | SU HOSPITALS OR “MEDICAL hospitals” OR “HEALTH facilities” OR “HOSPITAL administration” OR “HEALTH facility administration” OR “HEALTH services administration” OR “BUSINESS management of hospitals” OR “BUSINESS management of health facilities” |
| S5 | TI ( hospital OR hospitals ) OR AB ( hospital OR hospitals ) OR KW ( hospital OR hospitals ) |

| | |
|---|---|
| S6 | TI ( (health or healthcare) N3 (facility or facilities) ) OR AB ( (health or healthcare) N3 (facility or facilities) ) OR KW ( (health or healthcare) N3 (facility or facilities) ) |
| S7 | S4 OR S5 OR S6 |
| S8 | TI "rural referral center*" OR AB "rural referral center*" OR KW "rural referral center*" |
| S9 | S3 AND S7 |
| S10 | S8 OR S9 |
| S11 | SU "HOSPITAL mergers" |
| S12 | TI ( merg* or consolidat* or closure* or close or closes or closed or closing or acquisition* or acquir* ) OR AB ( merg* or consolidat* or closure* or close or closes or closed or closing or acquisition* or acquir* ) OR KW ( merg* or consolidat* or closure* or close or closes or closed or closing or acquisition* or acquir* ) |
| S13 | S11 OR S12 |
| S14 | S10 AND S13 |
| S15 | S14 + Limiters - Published Date: 20100101-; Language: English<br>Search modes - Boolean/Phrase |

### List of US Rural Health Grey Literature Sources

| **Grey Literature Databases** | | |
|---|---|---|
| NY Academy of Medicine Grey Literature Report | ProQuest Dissertations and Theses | Ideas Database (Federal Reserve Bank of St. Louis Economic Research Division) |
| **Preprint Servers** | | |
| medRxiv | OSF Preprints | |
| **Search Engines** | | |
| Google | Google Scholar | |
| **FOHRP-Funded Research Centers (past and present)** | | |
| NORC Walsh Center for Rural Health Analysis | North Dakota and NORC Rural Health Reform Policy Research Center | Rural Telehealth Research Center |

| | | |
|---|---|---|
| Upper Midwest Rural Health Research Center | West Virginia Rural Health Research Center | WICHE Center for Rural Mental Health Research |
| Maine Rural Health Research Center | North Carolina Rural Health Research and Policy Analysis Center | RUPRI Center for Rural Health Policy Analysis |
| Rural and Underserved Health Research Center | Rural Health Equity Research Center | Rural and Minority Health Research Center |
| Southwest Rural Health Research Center | University of Minnesota Rural Health Research Center | WWAMI Rural Health Research Center |
| **Policy and Research Analysis Initiatives** | | |
| Center for Economic Analysis of Rural Health | Rapid Response to Requests for Rural Data Analysis | RUPRI Health Panel: Rural Policy Analysis and Applications |
| **Government Agencies and Departments** | | |
| USA.gov | Centers for Disease Control and Prevention | Government Accountability Office |
| United States Department of Agriculture | Health Resources & Services Association | Federal Office of Rural Health Policy |
| Centers for Medicare and Medicaid Services | US Department of Health and Human Services | National Institutes of Health |
| Agency for Healthcare Research and Quality | National Conference of State Legislatures | State Offices of Rural Health |
| **Foundations, Think Tanks, Associations** | | |
| Think Tank Search | Robert Wood Johnson Foundation | Roosevelt Institute |
| Kaiser Family Foundation | Patient Centered Outcomes Research Institute | Commonwealth Fund |
| Ford Foundation | Pew Research Center | Pew Trusts |
| Advisory Board | National Organization of State Offices of Rural Health | Rural Health Information Hub |
| Rural Health Research Gateway | The Aspen Institute | Brookings |
| Mercatus Center | Center on Budget and Policy Priorities | Rand Corporation |
| National Bureau of Economic Research | Bipartisan Policy Center | Center for American Progress |
| National Rural Health Association | American Hospital Association | National Academies Press |
| **State Rural Health Associations** – by state | | |

### Study Selection and Data Extraction Processes

Records identified during the search were deduplicated using Zotero(Roy Rosenzweig Center for History and New Media, 2022) Duplicates Merger plug-in.(Frangoudes, 2021/2022) Results were imported into Covidence(Veritas Health Innovation, 2022) software and remaining duplicates were removed. Articles were included if they 1) discussed any outcomes or impacts of hospital closures and/or mergers, and 2) either focused on rural hospitals or outcomes for rural hospitals were reported separately from urban hospitals. Following pilot tests of the screening criteria with three of the authors (AC, KS, AG), both title and abstract screening and full-text screening were conducted independently by two of the authors (AC, KS) with disagreements resolved through discussion and consensus. Articles were excluded from this pool if all the data analyzed were collected prior to 2010, if the article reported only on data from a country other than the USA, or if the article was published in a language other than English. Articles were not excluded based on study design or if they reported on non-empirical work (e.g., commentaries, editorials, conceptual papers, etc.).

### Title and Abstract Screening

In this phase of screening, we examine all of the titles and abstracts imported from our search into Covidence.

#### *Screening Criteria*

In order to INCLUDE an article, it must satisfy ALL inclusion criteria and NONE of the exclusion criteria.

##### Exclusion

- Title/Abstract not in English
- Title/Abstract explicitly states it reports only on a country other than the US
- Title/Abstract explicitly states that data reported are from pre-2010

##### Inclusion

- Title/Abstract refers to rural hospitals
- Title/Abstract refers to a closure or a merger of at least one hospital with another health entity

#### *Screening Procedure*

- View the abstract of the record presented to you in Covidence
  - If there is no abstract, see TITLE ONLY screening criteria
- Quickly scan the title and abstract and EXCLUDE if:
  - Abstract is not in English
  - Article is explicitly reporting on a country other than the USA
  - Abstract explicitly states that data are from pre-2010

If not excluded yet, then the abstract needs a deeper read

- Does the abstract refer to rural hospitals?
  - To answer yes, the abstract must use terms that refer to rural environment, to specific rural areas in the USA, and/or specific rural hospital designations

| Rural Terms | Rural Regions | Rural Hospital Designations |
|---|---|---|
| • Rural<br>• Remote<br>• Frontier<br>• Isolated<br>• Non-core<br>• Non-metro<br>• Other | • Appalachia<br>• Delta Region<br>• Adirondack<br>• Ozarks<br>• (Indian, native American, other term) reservation<br>• Other | • Critical access hospital (CAH)<br>• Disproportionate Share Hospital (DSH)<br>• Sole Community Hospital (SCH)<br>• Low-Volume Hospital (LVH)<br>• Medicare Dependent Hospital (MDH)<br>• Tribal or Indian Health Services (HIS) Hospital<br>• Rural Referral Center<br>• Other |

- Does the abstract refer to a closure or merger of at least one hospital with another health entity?

| Terms | Caveats |
|---|---|
| • Merge, merger | |
| • Acquisition, acquire | *may see discussions of hospital- or community-acquired conditions– these are not relevant |
| • Affiliation, affiliate (especially if talking about health network or health system affiliation) | *may see this term used for a physician affiliation with a hospital – this is not relevant<br>*may see this term used for a hospital affiliated with a religious faction (eg. Catholic hospitals) – these *may* not be relevant, but they also could be – be careful! |
| • Consolidation | |

| | |
|---|---|
| • Closure, closing | |
| • Conversion | Hospitals that convert into other types of facilities are **included in this search** |
| • Integration | If referring to integration of a hospital within a system or network |

#### *TITLE ONLY Screening Procedure*

- If there is no abstract presented, screen the entry based on the title alone using the following abbreviated criteria:
- EXCLUDE if:
    - Title is not in English
    - Title is explicitly reporting on a country other than the USA
    - Title explicitly states that data are from pre-2010
- Does the title refer to rural hospitals?
- Does the title refer to a closure or merger of at least one hospital with another health entity?

#### *Pilot Test of Screening Criteria*

During an initial meeting, the project lead will introduce and explain the various inclusion and exclusion criteria and how to apply them for maximum concordance.

A subset of 25 abstracts have been selected from the search results by the project lead. These articles were selected by sorting the results by “recent” in Covidence and selecting the first page of results. We note that the selected subset contains a higher proportion of articles that will be “included” compared to the general search results, but that this was deemed necessary in order to properly test the inclusion and exclusion criteria. A random selection was initially drawn from the search results using a random number generator, but the subset contained zero includable items.

Each screener will independently screen the 25 abstracts in the subset and record their screening responses on the screening pilot form.

Once the sample abstracts have been reviewed by all screeners, the project lead will test concordance between responses and calculate inter-rater agreement. If inter-rater agreement (Kappa value) is less than 0.61 (“substantial” agreement), the screening team will meet to discuss screening criteria requiring clarification or refinement. Subsequent pilot tests will continue the same way until an adequate inter-rater agreement is reached.

#### *Title and Abstract Screening Cheat Sheet*

| | | | |
|---|---|---|---|
| **Scan the Title and Abstract** | | | |
| Abstract is not in English | | | **EXCLUDE** |
| Article is explicitly reporting on a country other than the USA | | | |
| Abstract explicitly states that data are from pre-2010 | | | |
| **Deeper Read of Title and Abstract** | | | |
| Does the abstract refer | **Rural Terms:**<br>• Rural | • Isolated<br>• Non-core | **I N** |

| | | | |
|---|---|---|---|
| to rural hospitals? | • Remote<br>• Frontier | • Non-metro<br>• Other | C<br>L<br>U<br>D<br>E |
| | **Rural Regions:**<br>• Appalachia<br>• Delta Region<br>• Adirondack | • Ozarks<br>• (Indian, Native American, other term) reservation<br>• Other | |
| | **Rural Hospital Designations**<br>• Critical access hospital (CAH)<br>• Disproportionate Share Hospital (DSH)<br>• Sole Community Hospital (SCH) | • Low-Volume Hospital (LVH)Medicare Dependent Hospital (MDH)<br>• Tribal or Indian Health Services (HIS) Hospital<br>• Rural Referral Center<br>• Other | |
| Does the abstract refer to a closure or merger of at least one hospital with another health entity? | Merge, merger | | |
| | 1. Acquisition, acquire | *hospital- or community-acquired conditions are not relevant | |
| | 2. Affiliation, affiliate<br><br>(especially health network or health system affiliation) | physician affiliation with a hospital – not relevant<br><br>*hospital affiliated with a religious faction (eg.Catholic hospitals) –*may* not be relevant, but they also could be – be careful! | |
| | 3. Consolidation | | |
| | 4. Closure, closing | | |
| | 5. Conversion | Hospitals that convert into other types of facilities are **included** in this search | |
| | 6. Integration | If referring to integration of a hospital within a system or network | |

*Pilot Title/Abstract Screening Form (Sample)*

Please use this form to complete your pilot set of abstracts.
Email*
Valid email

Screener Name
Short answer text

#2172 - Herman 2014

Is the abstract in a language other than English?
*

Option 1

No

Does it explicitly report on a country other than the USA?
*

Yes

No

Does it explicitly state that ALL data are from pre-2010?
*

Yes

No

Does the abstract refer to rural hospitals?
*

Yes

No

Does the abstract refer to a closure or merger of at least one hospital with another health entity?
*

Yes

No

Does the abstract refer to impacts or outcomes of merger or closure?

*

Yes

No

***OVERALL ASSESSMENT***
*

INCLUDE

EXCLUDE

Comments or questions
Long answer text

### Full-Text Screening

This phase of screening is where we refine our included articles to only collect those that help to answer the research question:

**What is known from the literature about the effects of rural hospital closures and mergers in the Affordable Care Act era?**

The key word to focus on here is EFFECTS. We would like to know what happens AFTER a hospital closes or merges with another health entity. We want to exclude papers that exclusively focus on merger or closure AS THE OUTCOME event.

#### *Screening Criteria*

In order to INCLUDE an article, it must satisfy ALL inclusion criteria and NONE of the exclusion criteria.

#### *Exclusion*

- *IF REPORTING ON RESEARCH: Data used in the study are from prior to 2010.*
    - In the introduction and/or the methods, determine the date range for the data. If ALL of the data were collected in 2009 or earlier, EXCLUDE
- *Paper reports on data from a country other than the US*
    - EXCLUDE
- *Paper not in English*
    - EXCLUDE

#### Inclusion

- *Hospital closure and/or merger is **a main topic** of the article*
    - Paper doesn't just use rural hospital closures or mergers or system affiliation" to frame or bracket the research
        - Explanation: Many articles include discussions of rural hospital closures in their introductions and/or conclusions to tie their work into the current literature.
        - Can recognize this when the discussion of rural hospital closures or mergers is superficial and limited to one or two sentences in the introduction and/or the conclusion.

      **OR**
    - **INCIDENCE** of merger/system/network affiliation (or synonym) or closure is considered as a variable in a study AND results report on this
        - In this case, the relevant text in the article <u>may be short and seem insubstantial</u>. Please INCLUDE
- *Rural hospitals are explicitly discussed in relation to the hospital closure/merger*
    - Rural hospitals are the sole focus of the article

      **OR**
    - Merger/closure outcomes related to rural hospitals are reported separately from urban hospitals
- *The article discusses one or more **outcomes/effects** of rural hospital closure and/or merger*
    - In a research study, the outcomes reported are measured <u>after</u> hospital closure or merger has occurred (note: may be a before/after, but after MUST be reported)
    - Closure or merger <u>may not</u> be discussed solely **as the outcome** of something else

#### *Screening Procedure*

- View the full text linked in Covidence
- Quickly scan the article and EXCLUDE if:
  - Article is not in English
  - Article is explicitly reporting on a country other than the USA
  - Article explicitly states that data are from pre-2010

If not excluded yet, then the paper needs a deeper read

- *Hospital closure and/or merger is* ***a main topic*** *of the article*
  - Paper doesn't just use rural hospital closures or mergers to frame or bracket the research
    **OR**
  - **INCIDENCE** of merger/system/network affiliation (or synonym) or closure is considered as a variable in a study AND results report on this

  *Note that department or service line closures ALONE do not meet the criteria

| **Terms** | **Caveats** |
|---|---|
| • Merge, merger | |
| • Acquisition, acquire | *may see discussions of hospital- or community-acquired conditions– these are not relevant |
| • Affiliation, affiliate (especially if talking about health network or health system affiliation) | *may see this term used for a physician affiliation with a hospital – this is not relevant<br>*may see this term used for a hospital affiliated with a religious faction (eg. Catholic hospitals) – these *may* not be relevant, but they also could be – be careful! |
| • Consolidation | |
| • Closure, closing | |
| • Conversion | Hospitals that convert into other types of facilities are **included in this search** |
| • Integration | If referring to integration of a hospital within a system or network |

IF YES, CONTINUE:

- *The merger/closure of RURAL hospitals is explicitly discussed*
  - Rural hospitals are the sole focus of the article and merger/closure is discussed
    **OR**
  - Merger/closure outcomes related to rural hospitals are <u>reported separately</u> from urban hospitals

| **Rural Terms** | **Rural Regions** | **Rural Hospital Designations** |
|---|---|---|

| | | |
|---|---|---|
| • Rural<br>• Remote<br>• Frontier<br>• Isolated<br>• Non-core<br>• Non-metro<br>• Other | • Appalachia<br>• Delta Region<br>• Adirondack<br>• Ozarks<br>• (Indian, native American, other term) reservation<br>• Other | • Critical access hospital (CAH)<br>• Disproportionate Share Hospital (DSH)<br>• Sole Community Hospital (SCH)<br>• Low-Volume Hospital (LVH)<br>• Medicare Dependent Hospital (MDH)<br>• Tribal or Indian Health Services (HIS) Hospital<br>• Rural Referral Center; other |

IF YES, CONTINUE

- *The article discusses one or more* ***outcomes/effects*** *of rural hospital closure and/or merger*
    - In a research study, the outcomes reported are measured after hospital closure or merger has occurred (note: may be a before/after, but after MUST be reported)
    - Closure or merger may not be discussed solely **as the outcome** of something else

IF YES, INCLUDE

#### Exclusion Reasons

When you click EXCLUDE in Covidence, you are asked to provide a reason. Please indicate the FIRST reason for exclusion, working down the cheat sheet in order.

- Article is not in English
- Article is about a country other than the USA
- Data reported are ALL from pre-2010
- Hospital closure/merger is not a main topic
- Rural hospitals are not explicitly discussed in relation to the hospital closure/merger
- Merger or closure is only discussed as an OUTCOME

#### *Pilot Test of Full-Text Criteria*

During an initial meeting, the project lead will introduce and explain the various inclusion and exclusion criteria and how to apply them for maximum concordance.

A subset of 5 articles has been selected from the full-text screening set by the project lead to be used in inclusion/exclusion criteria instruction. These articles were selected to illustrate particular aspects of the screening criteria. These 5 articles will be screened together in the initial meeting.

A further subset of 10 articles has been selected by the project lead from the full-text screening set. Pilot screeners will independently complete these in two rounds of 5 articles, recording their responses on a screening form. Feedback and/or discussion will be scheduled as needed after each round.

Once the sample abstracts have been reviewed by all screeners, the project lead will test screener concordance after each round of screening and calculate inter-rater agreement. If inter-rater agreement (Kappa value) is less than 0.61 (“substantial”), the screening team will meet to discuss screening criteria requiring clarification or refinement. (If Cohen’s Kappa value cannot be calculated, and agreement of >80% is required.) Subsequent pilot tests will continue the same way until satisfactory inter-rater agreement is reached.

#### *Full-Text Screening Cheat Sheet*

<table>
<tr><td colspan="3">1. Scan the Article</td></tr>
<tr><td colspan="2">Article is not in English</td><td rowspan="3">If any,<br>EXCLUDE</td></tr>
<tr><td colspan="2">Article is explicitly reporting on a country other than the USA</td></tr>
<tr><td colspan="2">Article explicitly states that ALL data are from pre-2010</td></tr>
<tr><td colspan="3">2. Deeper Read of Article</td></tr>
<tr><td rowspan="3">Hospital closure and/or merger is a main topic of the article</td><td>• Paper doesn’t just use rural hospital closures or mergers to frame or bracket the research</td><td rowspan="8">If all,<br><br>I<br>N<br>C<br>L<br>U<br>D<br>E</td></tr>
<tr><td>OR</td></tr>
<tr><td>• INCIDENCE of merger/system/network affiliation (or synonym) or closure is considered as a variable in a study AND results report on this</td></tr>
<tr><td rowspan="3">The merger/closure of RURAL hospitals is discussed</td><td>• Rural hospitals are the sole focus of the article and merger/closure is discussed</td></tr>
<tr><td>OR</td></tr>
<tr><td>• Merger/closure outcomes related to rural hospitals are <u>reported separately</u> from urban hospitals, with or without discussion</td></tr>
<tr><td rowspan="2">The article discusses one or more outcomes/ effects of rural hospital closure and/or merger</td><td>• In a research study, the outcomes reported are measured <u>after</u> hospital closure or merger has occurred (note: may be a before/after, but after MUST be reported)</td></tr>
<tr><td>• Closure or merger <u>may not</u> be discussed solely as the outcome of something else</td></tr>
</table>

Note: When you click EXCLUDE in Covidence, you are asked to provide a reason. Please indicate the FIRST reason for exclusion, working down the cheat sheet in order.

- Article is not in English
- Article is about a country other than the USA
- Data reported are ALL from pre-2010
- Hospital closure/merger is not a main topic
- Rural hospitals are not explicitly discussed
- Merger or closure is only discussed as an OUTCOME

#### *Pilot Full-Text Screening Form (Sample)*

Full Text Screening Form

- Please use this form to complete your pilot set of abstracts.
- Please fill the form out once for each article (you won't see all the articles listed here the way we did for the Ti/Ab screening so that you can stop and start as needed).

Screener Name

Your answer

Article Number (from the filename)

Your answer

Is the article in a language other than English?
Yes/No

Does it explicitly report on a country other than the USA?
Yes/No

Does it explicitly state that ALL data are from pre-2010?
Yes/No

Is hospital closure and/or merger a **main topic** of the article?
Yes/No

Rural hospitals are explicitly discussed?
Yes/No

The article discusses one or more **outcomes/effects** of rural hospital closure or merger?
Yes/No

Final Decision
Include/Exclude

Reason for Exclusion
Choose

- Article is not in English
- Article is about a country other than the USA
- Data reported are ALL from pre-2010
- Hospital closure/merger is not a main topic
- Rural hospitals are not explicitly discussed
- Merger or closure is only discussed as an OUTCOME

#### Data Extraction and Analysis

Data were charted from the included articles by the author according to a piloted extraction framework. For records which reported the results of research studies related to closure or merger outcomes (“primary studies”), we collected information about the study objectives, methods, design, data sources, and outcomes. For records reporting on knowledge syntheses, we collected information about the review objectives, methods, data sources, and findings. In this paper, we report on primary studies; our analysis of non-study records will be examined in a future publication.

### Extraction Form Development

Prior to the full-text screening stage, a preliminary data extraction form was created in Google Forms to capture content from two categories of records:

- Records reporting the results of a study
- Records not reporting the results of a study (but may be reporting on reports of studies)

The forms were revised on 11 July, 2023, after the extraction pilot test was completed but prior to beginning the extraction process.

#### *Section 1: All records bibliographic data*

This section is to be completed for all records.

| **Data collection field** | **Description** | **Answer type** | **Answer options** |
|---|---|---|---|
| CovidenceID | ID number assigned to the record in Covidence. Found by looking the article up in Covidence. | Short answer | n/a |
| Lead Author Name | Full name of lead author as listed in the publication header/byline | Short answer | n/a |
| Lead Author Affiliation | Affiliation of lead author as listed in the publication header/byline | Short answer | n/a |
| Reference | Copied from Covidence | Short Answer | n/a |
| Year of Publication | Year of publication as listed in the citation | Short answer | n/a |
| Publication Title | Title of the publication in which the record appears | Short answer | n/a |
| Publication Audience | Intended audience of publication (if this is evident from the text, source, and/or journal) | Multiple select | • Academic<br>• Clinical<br>• Management practitioners<br>• Policymakers<br>• Unclear<br>• Other? |
| Type of Record | Type of record | Single select | • Academic journal article<br>• News release/news article |

| | | | |
|---|---|---|---|
| | | | • Podcast<br>• Blog/website<br>• Presentation/Poster/ Webinar<br>• Policy/Issue Brief/ White Paper<br>• Dissertation<br>• Other? |
| Does this report on the results of a research study? | Indicate whether the record reports the results of research | Yes/No | • Yes<br>• no |

If the record reports the results of a research study, complete section 2. (Note: if a record contains a description of methods used, it is likely a research study)

If the record does not report the results of a research study, complete section 3. Note that if an article reports on and references the primary source of that research, it does NOT report on a research study directly.

#### *Section 2: Information about Reported Study*

This section is completed for any records that report the results of a research study in any format/medium. Where possible, the authors' words are captured using quotation marks and page numbers to identify direct quotes.

| **Data collection field** | **Description** | **Answer type** | **Answer options** |
|---|---|---|---|
| Objective/research question | The objective or research question, if possible, in the authors' words (use quotations and page numbers). In a case where the objective or question is said differently in various places, select the "best" | Long answer | n/a |
| Research methodology | Categorize the research by broad research methodology. | Single select | • Qualitative<br>• Quantitative<br>• Mixed Methods<br>• Research Synthesis<br>• Other<br>• Unclear |
| Brief discussion of research design/methods | Where possible, select the authors' words (in quotations | Long answer | n/a |

| | | | |
|---|---|---|---|
| | with page numbers) describing the research approach in terms of study design and/or methods. If not stated, leave blank. | | |
| Object of study | What is being studied by the researchers?<br>*In quantitative research, these would be the primary independent variables. If mergers/closures are only used as secondary independent variables, "something else" is selected. If merger or closure is only used as an outcome variable, the record should be returned to screening.* | Single select | • Hospital closures<br>• Hospital mergers<br>• Both hospital closures and mergers<br>• Something else |
| Unit of analysis for impacts/outcomes measured? | Identify at which level outcomes or impacts are **measured** | Multiple select | • Individual/patient<br>• Community - geographic (eg. town, village, neighborhood...)<br>• Community - social (eg. migrant workers, social identities...)<br>• Hospital<br>• Health Network<br>• County<br>• Other |
| Unit of analysis for impacts/outcomes reported? | Identify at which level outcomes or impacts are **reported** | Multiple select | • Individual/patient<br>• Community - geographic (eg. town, village, neighborhood...) |

| | | | |
|---|---|---|---|
| | | | • Community - social (eg. migrant workers, social identities...)<br>• Hospital<br>• Health Network<br>• County<br>• Other |
| List the geography/ies covered | If National, state "National". For states, use two-letter abbreviation) | Short answer text | n/a |
| Data collection period – Merger or closure | Identify the years in which the mergers and/or closures occurred. *If no merger or closure outcome data collected, use "N/A".* | Short answer text | n/a |
| Data collection period - Outcomes | Identify the years in which the study outcome data were collected. *If no merger or closure outcome data collected, use "N/A".* | Short answer text | n/a |
| Definition of Rural | Capture the definition or classification of rurality used to determine rural/urban status. Be sure to capture the definition/ classification as well as any thresholds or cut-points applied. Indicate page numbers if possible. *If the authors do not report, note "not reported". If rural* | Long answer text | n/a |

| | | | |
|---|---|---|---|
| | *classification does not apply, note "N/A"* | | |
| Did the authors use an existing data set? | Note if the authors used sources of existing data in their study. | Yes/no | Yes/no |
| If yes, provide details of the data sources used in the study | Describe all data sources used in the study (use authors' words where possible, paraphrase as necessary). *For research using existing datasets, capture as much detail as necessary to further research the repository (reference page number).* | Long answer text | n/a |
| Did the authors collect primary data? | Note if the authors used collected primary data in their study. | Yes/no | Yes/no |
| If yes, describe the sample and reference page numbers | *For research employing primary data collection (quantitative, qualitative, survey…), describe the sample and reference the page number. If a validated data collection tool is used (eg. a common survey), indicate that here.* | Long answer text | n/a |
| Outcomes/impacts referenced in introduction/background and discussion sections | Capture text from the non-study data that describes | Long answer text | n/a |

| | | | |
|---|---|---|---|
| | outcomes or impacts of mergers or closures. Use the authors' words where possible. If referenced/cited, capture the citation. | | |
| Outcome/Impact measures | Capture the outcome or impacts measured in the study or emerging from the analysis, including the definition of the outcome in the authors' words, where possible. (If no merger or closure outcome data collected, use N/A) | Long answer text | n/a |
| Outcome/Impact findings | Summarize the findings and note page numbers (in the same order listed for measures) (If no merger or closure outcome data collected, use N/A) | Long answer | n/a |
| Research Paradigm | What is the research paradigm leveraged in the current research? Where stated, use the authors' description of their paradigm.<br><br>*Capture only if explicitly stated by the authors.* | Single select | • Positivism<br>• Constructivism/interpretivism<br>• Pragmatism<br>• Subjectivism<br>• Critical<br>• Not stated<br>• Other |

#### *Section 3: Information about Non-Research Record*

This section is completed for any records that DO NOT report the direct results of a research study.

| Data collection field | Description | Answer type | Answer options |
|---|---|---|---|
| Reports on a particular research study published elsewhere? | Indicate whether the record showcases, publicizes, or in any way disseminates a given research study's results. *If the record references several studies but does not focus on one, answer "no" here.* | Yes/No | • Yes<br>• No |
| If yes, note the reference | Provide the link or reference as provided in the record. If not applicable, leave blank. | Long answer text | n/a |
| Source(s) of Data | Describe all data sources used in the record (use authors' words where possible, paraphrase as necessary). *If no data sources used in record, state "N/A". If unclear, state "unclear"* | Long answer text | n/a |
| Article main topic | What is being reported on in this record? | Single select | • Hospital closures<br>• Hospital mergers<br>• Both hospital closures and mergers<br>• Something else |
| Outcomes of closure/merger discussed | Capture the reported impacts and/or outcomes of closure or merger. Provide authors' words where possible with page numbers | Long answer | n/a |
| Unit of analysis for impacts/outcomes? | Identify at which level outcomes or impacts are measured or reported | Multiple select | • Individual/patient<br>• Community - geographic (eg. town, village, neighborhood...)<br>• Community - social (eg. migrant workers, social identities...)<br>• Hospital<br>• Health Network<br>• County<br>• Unclear<br>• Not applicable |

| | | | |
|---|---|---|---|
| | | | • Other? |
| List the geography/ies covered | If National, state “National”. For states, use two-letter abbreviation) | Short answer text | • n/a |

### Scoping Review Results

#### Overall Summary of Included Studies

Our search yielded 3697 records from the Medline, Scopus, and Business Source Complete databases, of which 1294 were duplicates and removed prior to screening. Title and abstract screening resulted in the exclusion of 2203 records. Full-text documents were retrieved and screened for 199 records from databases (one full text was unavailable). The gray literature search identified 333 records, of which 88 were duplicates and one was unavailable in full text. Of the 244 records grey literature reviewed in full, 97 were excluded. Following the full-text screening, 221 records were included in the review: 74 from databases and 147 from grey literature. Finally, for the purpose of this article, we excluded the 164 non-study records, resulting in 57 included records. A detailed PRISMA diagram is included on p. 21 of this supplement.

Of the 57 included records, there were 46 primary studies and two research syntheses. In cases where multiple reports were generated for one study (e.g., dissertation, journal article, policy brief, etc.), we used the peer-reviewed publication as the primary or definitive record, if applicable, or another publicly accessible report, such as a working paper, white paper, or brief.

Thirty-six (78.3%) of included studies were about rural hospital closures; the remaining 10 (21.7%) were about rural hospital mergers. Most studies used quantitative methods (n=37, 80.4%). Studies included administrative data (n=38, 82.6%), or primary data collection (n=10, 21.7%) and were either focused on the United States as a whole (n=31, 67.4%) or smaller geographies such as states or counties (n=15, 32.6%).

### Detailed Description of Included Studies - Closures

| ***Author, Year*** | ***Methodology (analysis)*** | ***Geographic Scope*** | ***Period of Study*** | ***Rural Definition (threshold)*** | ***Data Sources (purpose)*** | ***Outcomes, Impacts Measured (significant finding)*** |
|---|---|---|---|---|---|---|
| *Alexander & Richards 2023*<br><br>*(D. E. Alexander & Richards, 2021; D. Alexander & Richards, 2023)* | Quantitative (difference in differences) | National | 2001-2019 | Sheps Center Definition | • Sheps Center Rural Hospital Closure File (identify closed hospitals)<br>• AHA Annual Survey 2005 (identification of control hospitals)<br>• BLS QCEW 2001-2018 (outcome data)<br>• Federal Reserve Bank of New York Consumer Credit Panel/Equifax (outcome data)<br>• Home Mortgage Disclosure Act Database (outcome data) | *Employment/Workforce*<br>• Total employment - all, private sector, health care, non-health care (decreased for all, healthcare, private sector)<br>• Neighboring county employment<br>*Financial*<br>• Consumer credit risk score<br>• Consumer total balance past due<br>• Consumer past-due debts<br>• Consumer bankruptcies<br>*Well-being*<br>• Mortgage loans denied, originated, purchased<br>• Mortgage loan amount<br>• Mortgage applicant income |

| *Author, Year* | *Methodology (analysis)* | *Geographic Scope* | *Period of Study* | *Rural Definition (threshold)* | *Data Sources (purpose)* | *Outcomes, Impacts Measured (significant finding)* |
|---|---|---|---|---|---|---|
| *Andreyeva et al. 2022*<br><br>*(Andreyeva et al., 2022)* | Quantitative (difference in differences) | State | 2014-2019 | RUCC 2013 (4-9) | • CMS POS 2015-2018 (identify closed hospitals)<br>• AHA Annual Survey (identify closed hospitals)<br>• American Hospital directory (identify closed hospitals)<br>• Claims data from large commercial insurer (measure outcomes)<br>• Dartmouth Atlas HSA crosswalk (derive Herfindahl-Hirschman index for covariate analysis)<br>• ACS 5-year data (community descriptive data for covariate analysis) | *Utilization*<br>• Visits – outpatient, ED, inpatient, ED with admission, specialist, primary care (decrease for outpatient, ED)<br>*Financial*<br>• PMPM spending - outpatient, ED, inpatient, ED with admission, specialist, primary care |
| *Bell et al. 2023*<br><br>*(Bell et al., 2022, 2023)* | Quantitative (matched case control, conditional fixed effects interrupted time series) | National | 2006-2018 | RUCA 2010 (4-6 = large rural, 7-9 = small rural, 10= isolated rural) | • Sheps Center Rural Hospital Closure File (identify closed hospitals)<br>• CMS POS (measure outcome variables)<br>• American Communities Survey (covariate analysis) | *Access to Facilities and Services*<br>• Proportion of rural zip codes within 10 miles of FQHC/CHC (increase) |
| *CapitalLink 2022*<br><br>*(Capital Link, 2022)* | Qualitative (key informant interviews) | Community | 2018 | n/a | • Interviews with senior leaders from a local CHC | *Well-being*<br>• Psychosocial (negative)<br>• Morale (negative)<br>*Access to Facilities and Services*<br>• Non-hospital services - FQHCs, pharmacy, other (increased) |

| ***Author, Year*** | ***Methodology (analysis)*** | ***Geographic Scope*** | ***Period of Study*** | ***Rural Definition (threshold)*** | ***Data Sources (purpose)*** | ***Outcomes, Impacts Measured (significant finding)*** |
|---|---|---|---|---|---|---|
| *Chatterjee et al. 2022*<br><br>*(Chatterjee et al., 2022)* | Quantitative (difference in differences) | National | 2005-2018 | CBSA (5, 6) IRR | • Sheps Center Rural Hospital Closure File (identify closed hospitals)<br>• BLS Bureau of Economic Analysis Quarterly Workforce Indicators (outcome data)<br>• Social Security Administration Disability Data (outcome data)<br>• US Federal Reserve Economic Data (outcome data)<br>• RAND Corporation state statistics database (outcome data)<br>• US Census Bureau 2000 (outcome data) | *Workforce*<br>• Unemployment rate<br>• Employment to population ratio<br>• Labor force participation/population<br>• Total jobs<br>• Total health care jobs (decreased)<br>*Financial*<br>• Income per capita<br>• Credit scores<br>• Bankruptcy filings<br>*Well-being*<br>• Disability program participation<br>• Population |
| *Chaudhary et al. 2019*<br><br>*(S. Chaudhary et al., 2019; Troske, 2018a, 2018b; Troske & Davis, 2019)* | Quantitative (difference in differences) | National | 2010-2015 | US Department of Agriculture UIC 2003 (rural - 4-9, wilderness - 7-12) | • Sheps Center Rural Hospital Closure File (identify closed hospitals)<br>• NEMSIS (outcome data)<br>• CMS Hospital Compare (identify control hospitals)<br>• CMS POS 2010-2016 (descriptive data) | *Utilization*<br>• Ambulance total call time (increased)<br>• Ambulance time-to-scene (decreased)<br>• Ambulance scene time<br>*Access to Facilities and Services*<br>• Ambulance - transport time (increased) |

| ***Author, Year*** | ***Methodology (analysis)*** | ***Geographic Scope*** | ***Period of Study*** | ***Rural Definition (threshold)*** | ***Data Sources (purpose)*** | ***Outcomes, Impacts Measured (significant finding)*** |
|---|---|---|---|---|---|---|
| *Durrance et al. 2023*<br><br>*(Durrance et al., 2024)* | Quantitative (difference in differences, event study) | National | 2005-2019 | RUCC (moderately rural = 4-7, most rural = 8-9) | • Sheps Center Rural Hospital Closure File (identify closed hospitals)<br>• National Vital Statistics System 2005-2019 (outcome data) | *Access to Facilities and Services*<br>• Births occurring in county of residence (decreased)<br>• Prenatal care initiation (increased for moderately rural)<br>*Well-being*<br>• Low birth weight (increased for moderately rural)<br>• Preterm births (increased for most rural)<br>• Apgar scores (decreased for moderately rural)<br>• Infant mortality<br>*Utilization*<br>• Cesarian delivery (increased for moderately rural)<br>• Likelihood of induction<br>• Birth in hospital (increased for moderately rural, most rural) |
| *Edmiston 2019*<br><br>*(Edmiston, 2019)* | Quantitative | National | 2009-2018 | Other | • AHA Hospital Directory (identify closed hospitals)<br>• CMS HCUP (descriptive data)<br>• BLS QCEW (outcome data) | • Total employment<br>• Aggregate wages |

| Author, Year | Methodology (analysis) | Geographic Scope | Period of Study | Rural Definition (threshold) | Data Sources (purpose) | Outcomes, Impacts Measured (significant finding) |
|---|---|---|---|---|---|---|
| *Eilrich et al. 2015*<br><br>*(Eilrich et al., 2015)* | Mixed/multiple (quantitative - IMPLAN analysis, case study) | 16 communities in 13 states | unclear | RUCA (7-9) | • Sheps Center Rural Hospital Closure File (identify closed hospitals)<br>• CMS Medicare Cost Report and Claims Data (outcome data)<br>• Key informant interviews from one community and surrounding region (outcome data) | *Workforce*<br>• Employment - direct, total impact with multiplier<br>*Financial*<br>• Labor income - direct, total income with labor multiplier<br>• EMS operating costs (increase)<br>*Utilization*<br>• EMS transportation time and mileage (increase)<br>*Well-being*<br>• Community social well-being - morale (decrease) |

| | | | | | | |
|---|---|---|---|---|---|---|
| *GAO 2020*<br><br>*(US Government Accountability Office, 2020)* | Quantitative | National | 2012-2018 | FORHP definition | • Sheps Center Rural Hospital Closure File (identify closed hospitals)<br>• CMS HSAF (identify hospital service areas)<br>• HRSA AHRF 2021 and 2017 (outcome data<br>• CMS Medicare Variation files (outcome data)<br>• CMS POS files (outcome data)<br>• CMS Medicare HCUP (outcome data) | *Access to Facilities and Services*<br>• Number of healthcare providers per 100,000 population (lower for physicians, health care providers with some exceptions by physician specialty, higher for APRNs)<br>• Availability of hospital-based services (decreased except Substance Use Disorder treatment)<br>• Availability of non-hospital Medicare facilities – RHC, Community Mental Health Center, Ambulatory Surgical Center, FQHC<br>• Distance to nearest hospital with services (increased)<br>*Utilization*<br>• Number of inpatient stays (decreased)<br>• Number of outpatient visits (decreased)<br>• Number of ED visits (decreased)<br>• Number of hospital readmissions (smaller decrease)<br>*Well-being*<br>• Prevalence of chronic conditions among Medicare beneficiaries (higher)<br>*Financial* |

| *Author, Year* | *Methodology (analysis)* | *Geographic Scope* | *Period of Study* | *Rural Definition (threshold)* | *Data Sources (purpose)* | *Outcomes, Impacts Measured (significant finding)* |
|---|---|---|---|---|---|---|
| | | | | | | • Medicare spending on health care services (higher for inpatient services, higher increase for outpatient services) |
| *Gelbaugh 2021*<br><br>*(Gelbaugh & Advisory Board, 2021)* | Quantitative | National | 2017-2020 | RUCA (unknown) | • Sheps Center Rural Hospital Closure File (identify closed hospitals)<br>• US Census Bureau (delineate regions)<br>• Medicare Claims Data (outcome data) | *Utilization*<br>• Rates of ED utilization in Medicare beneficiaries |
| *Germack et al. 2019*<br><br>*(Germack et al., 2019)* | Quantitative (event study, difference in differences) | National | 1997-2016 | RUCC 2013 (4-9) | • HRSA Bureau of Health Workforce<br>• AHRF (identify counties with closures, outcome data) | *Access to Facilities and Services*<br>• Supply of Physicians (decrease for all physicians, primary care, and OB/Gyn) |
| *Germack et al. 2021*<br><br>*(Germack et al., 2021)* | Quantitative (event study, difference in differences) | National | 2010-2017 | RUCC 2013 (4-9) | • HRSA Bureau of Health Workforce<br>• AHRF (identify counties with closures, outcome data) | *Access to Facilities and Services*<br>• Supply of nurse practitioners (decrease at two years post-closure)<br>• Supply of Certified Registered Nurse Anesthetists |

| *Author, Year* | *Methodology (analysis)* | *Geographic Scope* | *Period of Study* | *Rural Definition (threshold)* | *Data Sources (purpose)* | *Outcomes, Impacts Measured (significant finding)* |
|---|---|---|---|---|---|---|
| *Gujral & Basu 2019*<br><br>*(Gujral & Basu, 2019)* | Quantitative (difference in differences) | 1 State | 1995-2011 | RUCA 2004 | • OSHPD discharge data (identify closed hospitals, outcome data)<br>• OSHPD hospital financial data (descriptive data)<br>• Dartmouth Atlas of Health Care crosswalks (identifying HSAs) | *Well-being*<br>• Inpatient mortality (increased)<br>*Utilization*<br>• Length of stay (increased)` |
| *Khushalani et al. 2023*<br><br>*(Khushalani et al., 2023)* | Quantitative (propensity score weighted regression and event study) | 9 States | 2010-2017 | Sheps Center Definition | • Sheps Center Rural Hospital Closure File (identify closed hospitals)<br>• HCUP State Inpatient Data (outcome data)<br>• Claritas PopFacts data (community descriptive data)<br>• Dartmouth Atlas HSA crosswalk (identify zip codes affected by closures) | *Utilization*<br>• Age-adjusted admission rates for ECSC<br>• Age adjusted admission rates for ACSC (higher)<br>• ALOS for ECSC (higher)<br>• ALOS for ACSC (lower)<br>*Well-being*<br>• In-hospital mortality for ECSC |

| *Author, Year* | *Methodology (analysis)* | *Geographic Scope* | *Period of Study* | *Rural Definition (threshold)* | *Data Sources (purpose)* | *Outcomes, Impacts Measured (significant finding)* |
|---|---|---|---|---|---|---|
| *Letheren et al. 2023*<br><br>*(Letheren et al., 2024)* | Qualitative (qualitative descriptive approach) | 1 Community | 2019-2020 | Sheps Center Definition | • Semi-structured telephone interviews with 24 adult (>30y) individuals from one community with a recent hospital closure | *Access to Facilities and Services*<br>• Travel time and distance to access care (increased)<br>• Transportation challenges<br>• Access to emergency and specialty care (decreased)<br>• Wait times to access services (increased)<br>*Workforce*<br>• Availability/supply of healthcare providers (decreased)<br>*Utilization*<br>• Workload/utilization of remaining health care providers (increased)<br>*Financial*<br>• Costs of hospital care - ability to pay, predictability of costs (decreased)<br>*Well-being*<br>• Familiar connections with health care providers (decreased) |

| *Author, Year* | *Methodology (analysis)* | *Geographic Scope* | *Period of Study* | *Rural Definition (threshold)* | *Data Sources (purpose)* | *Outcomes, Impacts Measured (significant finding)* |
|---|---|---|---|---|---|---|
| *Malone et al. 2022*<br><br>*(T. L. Malone et al., 2022)* | Quantitative (difference in differences) | National | 2001-2018 | OMB (non-metro) | • Sheps Center Rural Hospital Closure File (identify closed hospitals)<br>• IRS (outcome data)<br>• Census bureau (outcome data)<br>• BLS (outcome data) | *Financial*<br>• Annual income<br>*Workforce*<br>• Unemployment rate<br>• Labor force size (decreased for all hospital types)<br>*Health/Wellness/Quality*<br>• Population size (decreased for PPS hospital closures) |
| *Manlove & Whitacre 2017*<br><br>*(Manlove & Whitacre, 2017)* | Quantitative (econometric, difference in differences) | 20 States | 2012-2014 | Sheps Center Definition RUCC | • Sheps Center Rural Hospital Closure File (identify closed hospitals)<br>• ACS (descriptive and outcome data) | *Financial*<br>• Income (decreased relative to matched)<br>• Poverty level (increased relative to matched)<br>• Rent (decreased relative to matched)<br>• Home values<br>*Workforce*<br>• Unemployment (increased relative to matched)<br>• Travel time to work<br>• Work at home<br>• Occupational mix (decrease in construction relative to matched)<br>• Industry mix (decrease in some relative to nearest and matched) |

| *Author, Year* | *Methodology (analysis)* | *Geographic Scope* | *Period of Study* | *Rural Definition (threshold)* | *Data Sources (purpose)* | *Outcomes, Impacts Measured (significant finding)* |
|---|---|---|---|---|---|---|
| *McCarthy et al. 2020*<br><br>*(McCarthy et al., 2021)* | Quantitative (geospatial) | National | 2010-2016 | CBSA (micropolitan, non-metropolitan) Sheps Center definition of rural hospital | • Sheps Center Rural Hospital Closure File (identify closed hospitals)<br>• CMS POS files (geographic identification of hospitals)<br>• Census Data 2010 (geospatial population data) | *Access to Facilities and Services*<br>• Change in number of residents who can access a hospital within 15, 30, 45, 60 minutes (decreases in some geographies and thresholds)<br>• Change in proportion of residents who can access a hospital within 15, 30, 45, 60 minutes (decreases in some geographies and thresholds) |
| *MedPAC 2021*<br><br>*(Medicare Payment Advisory Commission (MEDPAC), 2021)* | Mixed/multiple (quantitative, survey, stakeholder interviews) | National | 2005-2018 | OMB UIC - (rural micropolitan, other rural) | • CMS Medicare Cost Report and Claims Data (outcome data)<br>• Interviews with stakeholders from three communities that experienced a recent hospital closure - community members, executives, clinician leaders (outcome data)<br>• Virtual site visits to three rural communities that experienced a recent hospital closure - included interviews with hospital executives, city/county government officials, clinician leaders, EMS | *Access to Facilities and Services*<br>• Access to non-hospital services (FQHCs, freestanding ED, urgent care, etc.) (increased)<br>• Travel to access care (increased)<br>• Reliance on EMS (increased)<br>• EMS transport times (increased)<br>*Utilization*<br>• Number of inpatient admissions, hospital outpatient visits (decreased)<br>• Number of evaluation and management visits across all settings (greater increase)<br>• EMS response times (decreased) |

| *Author, Year* | *Methodology (analysis)* | *Geographic Scope* | *Period of Study* | *Rural Definition (threshold)* | *Data Sources (purpose)* | *Outcomes, Impacts Measured (significant finding)* |
|---|---|---|---|---|---|---|
| *Merrell 2019*<br><br>*(Merrell, 2019)* | Quantitative (difference-in-difference-in-difference) | 2 states | 2010-2014 | Sheps Center Definition | • Sheps Center Rural Hospital Closure File (identify closed hospitals)<br>• US Census Bureau (identify matching counties)<br>• AHRQ HCUP SID and State Emergency Department Databases (outcome data) | *Health/Wellness/Quality*<br>• In-hospital mortality for ECSC (greater decrease)<br>• 30-day mortality |
| *Mike 2019*<br><br>*(Mike, 2020)* | Qualitative (hermeneutic phenomenology) | Region | 2013-2014 | US Census Bureau | • Qualitative interviews with eight African American older adults who had been treated for a previous emergency medical condition who reside in the region of a rural hospital closure | *Access to Facilities and Services*<br>• Access to healthcare treatment<br>• Increased distance to care<br>*Utilization*<br>• Friends and families supporting patient's health care needs |
| *Miller et al. 2020*<br><br>*(Miller et al., 2020)* | Quantitative (pre/post retrospective cohort with propensity matched control, difference-in-differences and quantile regression) | National | 2010-2016 | NEMSIS defined - based on 2003 UIC (rural, wilderness/frontier) | • CMS POS 2010-2016 (identify closed hospitals)<br>• AHRF 2010 (descriptive data)<br>• NEMSIS 2010-2016 (outcome data) | *Access to Facilities and Services*<br>• EMS response time<br>• EMS scene-to-patient time<br>• EMS activation time (increased)<br>• Transport time (increased) |

| *Author, Year* | *Methodology (analysis)* | *Geographic Scope* | *Period of Study* | *Rural Definition (threshold)* | *Data Sources (purpose)* | *Outcomes, Impacts Measured (significant finding)* |
|---|---|---|---|---|---|---|
| *Miller et al. 2021*<br><br>*(Miller et al., 2021)* | Quantitative (fixed effects and pooled generalized linear models) | National | 2006-2018 | OMB (non-metro) | • Sheps Center Rural Hospital Closure File (identify closed hospitals)<br>• CMS POS files (outcome data)<br>• Small Area Income and Poverty Estimates files 2006-2018 (descriptive data) | *Access to Facilities and Services*<br>• Probability of FQHC/RHC within 10 miles of a closed hospital (increase for FQHC) |
| *Mills 2022*<br><br>*(Mills, 2022)* | Quantitative (difference in differences) | National | 2005-2018 | Sheps Center Definition | • Sheps Center Rural Hospital Closure File (identify closed hospitals)<br>• Brown University LTCFocus Program (outcome data)<br>• AHRF (descriptive data) | *Access to Facilities and Services*<br>• Number of nursing homes (decreased)<br>• Number of nursing home beds (decreased) |
| *Mobley et al. 2020*<br><br>*(Mobley et al., 2020)* | Quantitative | National | 2009-2020 | Sheps Center Definition | • Sheps Center Rural Hospital Closure File (identify closed hospitals)<br>• National Provider Identifier data (outcome data) | *Access to Facilities and Services*<br>• Number of healthcare providers – Advanced Practice Providers, Primary Care Providers, specialty physicians |
| *Nikpay et al. 2021*<br><br>*(Nikpay et al., 2021)* | Quantitative (difference in differences) | National | 2012-2018 | RUCC (6-9) | • Sheps Center Rural Hospital Closure File (identify closed hospitals)<br>• CMS POS files (outcome data)<br>• CMS POS Medicare Public Use Files (outcome data) | *Access to Facilities and Services*<br>• EMS total miles (increased)<br>• EMS average miles (increased)<br>*Utilization*<br>• EMS total trips<br>• EMS proportion of non-emergent trips (increased) |

| *Author, Year* | *Methodology (analysis)* | *Geographic Scope* | *Period of Study* | *Rural Definition (threshold)* | *Data Sources (purpose)* | *Outcomes, Impacts Measured (significant finding)* |
|---|---|---|---|---|---|---|
| *Ramedani et al. 2022*<br><br>*(Ramedani et al., 2022)* | Quantitative (average rate of change) | National | 2003-2018 | Sheps Center Definition | • Sheps Center Rural Hospital Closure File (identify closed hospitals)<br>• AHA Annual Survey (outcome data) | *Utilization*<br>• Bystander hospital admissions (increased)<br>• Bystander hospital emergency department visits (increased)<br>*Financial*<br>• Bystander hospital profit margin<br>• Change in cost for bystander hospitals - nonprofit/for profit (increased) |
| *Smith JG et al. 2022*<br><br>*(J. G. Smith et al., 2022)* | Qualitative (thematic) | State | 2014-2020 | Other | • Qualitative interviews with 10 Registered Nurses or Licensed Vocational Nurses who worked at hospitals that closed between 2014-2020 in one Southwestern state | *Access to Facilities and Services*<br>• Patient access to care (negative)<br>• Patient outcomes (negative)<br>*Well-being*<br>• Social impacts (mixed)<br>• Community economic/employment (negative)<br>• Nurse psychosocial well-being (negative)<br>*Workforce*<br>• Job outcomes (negative) |
| *Smith TB et al. 2022*<br><br>*(T. B. Smith et al., 2022)* | Quantitative (linear regression, difference in differences) | 1 State | 2010-2021 | Sheps Center Definition | • Sheps Center Rural Hospital Closure File (identify closed hospitals)<br>• Alabama Department of Public Health EMS Database (outcome data) | • Total transport time (mixed effects by region) |

| ***Author, Year*** | ***Methodology (analysis)*** | ***Geographic Scope*** | ***Period of Study*** | ***Rural Definition (threshold)*** | ***Data Sources (purpose)*** | ***Outcomes, Impacts Measured (significant finding)*** |
|---|---|---|---|---|---|---|
| *Song & Saghafian 2019*<br><br>*(D. (Lina) Song, 2020; L. D. Song & Saghafian, 2019)* | Quantitative (difference in differences) | National | 2005-2015 | RUCA (greater than 4)<br>Critical Access Hospital designation | • Medicare Cost Reports and POS data (descriptive and outcome data)<br>• FFS Medicare inpatient claims (outcome data)<br>• Medicare beneficiary summary files (descriptive data)<br>• Hospital Compare (CMS) - Hospital Quality (outcome data)<br>• Hospital Consumer Assessment of Healthcare Providers and Systems (HCAHPS) (outcome data)<br>• AHRF (descriptive data)<br>• CMS state/county/plan enrollment data file (identification of study population) | *Financial*<br>• Nearby Hospital Operational efficiency (increased)<br>*Utilization*<br>• Nearby Hospital Bed utilization<br>• Nearby Hospital length of stay (decreased)<br>*Health/Wellness/Quality*<br>• Nearby Hospital 30-day readmissions<br>• Nearby Hospital 30-day mortality (increased) |

| ***Author, Year*** | ***Methodology (analysis)*** | ***Geographic Scope*** | ***Period of Study*** | ***Rural Definition (threshold)*** | ***Data Sources (purpose)*** | ***Outcomes, Impacts Measured (significant finding)*** |
|---|---|---|---|---|---|---|
| *Tennessee Health Care Campaign 2021*<br><br>*(Tennessee Health Care Campaign, 2021)* | Community Engaged Research (expert panel, listening sessions, key informant interviews) | State | 2018-2020 | not stated | • Expert panel of three physicians and practitioners who lived and worked in rural communities<br>• Five listening sessions with community leaders (patients, educators, elected officials, first responders, health care providers) from rural counties experiencing or at risk of hospital closure<br>• Key informant interviews with five leaders (hospital executives, physicians, EMS chief) in three rural locations where hospitals had closed | *Access to Facilities and Services*<br>• Access to specialists (decreased)<br>*Utilization*<br>• Stress on ambulance, other services (increased)<br>*Well-being*<br>• Psychosocial well-being (increased worry)<br>• Job creation (decreased)<br>• Ability to attract new residents to community (decreased)<br>*Financial*<br>• Business outcomes (decreased) |

| ***Author, Year*** | ***Methodology (analysis)*** | ***Geographic Scope*** | ***Period of Study*** | ***Rural Definition (threshold)*** | ***Data Sources (purpose)*** | ***Outcomes, Impacts Measured (significant finding)*** |
|---|---|---|---|---|---|---|
| *Thomas et al. 2015*<br><br>*(Thomas et al., 2015)* | Qualitative (non-random qualitative survey) | National | 2010-2014 | Other | • Qualitative surveys from 32 individuals from 22 hospitals in 13 different states | *Access to Facilities and Services*<br>• Access to health care services (decreased)<br>• Foregone care<br>• Loss of non-healthcare services<br>*Utilization*<br>• Stress on EMS providers (increased)<br>*Health/Wellness/Quality*<br>• Loss of large employer<br>• Community morale, identity (decreased)<br>• Economic growth (decreased)<br>*Financial*<br>• Burden of closed hospital debt<br>*Workforce*<br>• Job loss, unemployment rates (increased) |

| *Author, Year* | *Methodology (analysis)* | *Geographic Scope* | *Period of Study* | *Rural Definition (threshold)* | *Data Sources (purpose)* | *Outcomes, Impacts Measured (significant finding)* |
|---|---|---|---|---|---|---|
| *Vogler 2020*<br><br>*(Vogler, 2020a, 2020b)* | Quantitative (difference in differences) | National | 2003-2017 | Other | • Sheps Center Rural Hospital Closure File (identify closed hospitals)<br>• AHA Annual Survey Database (identify closed hospitals and descriptive data)<br>• QCEW (outcome data)<br>• BLS Local Area Unemployment Statistics (outcome data)<br>• IRS Statistics of Income (outcome data)<br>• Zillow Home Value Index (outcome data)<br>• US Housing and Urban Development (outcome data) | *Workforce*<br>• Employment (decreased)<br>• Unemployment rate<br>• Labor force participation (decreased) Private service sector employment (decreased)<br>• Non-hospital employment (decreased)<br>• Private goods-producing sector employment<br>*Financial*<br>• Per capita income (decreased)<br>• Median rents (decreased)<br>*Well-being*<br>• Total establishments<br>• Total population (decreased)<br>• Youth population (decreased)<br>• Adult population (decreased)<br>• Older adult population (decreased)<br>• Non-hospital establishments<br>• Private service sector establishments<br>• Private goods-producing sector establishments |

| *Author, Year* | *Methodology (analysis)* | *Geographic Scope* | *Period of Study* | *Rural Definition (threshold)* | *Data Sources (purpose)* | *Outcomes, Impacts Measured (significant finding)* |
|---|---|---|---|---|---|---|
| *Wishner et al. 2016*<br><br>*(Wishner et al., 2016)* | Qualitative (comparative case study) | Three Communities | 2015 | not stated | • Three cases of recent Prospective Payment System hospital closures, two in non-expansion states<br>• 6-8 community stakeholder interviews per case<br>• Publicly available documents related to closures<br>• State and regional analyses and planning initiatives<br>• Key informant/expert interviews | *Access to Facilities and Services*<br>• Access to care, emergency care, specialty care (decreased)<br>• Delayed/foregone care (increased)<br>• Transportation challenges (increased)<br>• Distance and travel time to access care (increased)<br>• Access to non-hospital services - e.g. FQHCs - (increased)<br>• Public sector services (decreased)<br>*Workforce*<br>• Supply of health care professionals (decreased)<br>• Jobs - non-health care (decreased)<br>*Well-being*<br>• Population size (decreased)<br>• Tax base (decreased)<br>• Ability to attract employers (decreased) |
| *Zahnd et al. 2023*<br><br>*(Zahnd et al., 2023)* | Quantitative (longitudinal analysis) | National | 2008-2017 | UIC 2013 (micropolitan, non-core) Critical Access Hospital designated hospitals | • Sheps Center Rural Hospital Closure File (identify closed hospitals)<br>• Dartmouth Atlas HSAs (define regions)<br>• AHA Annual Surveys (outcome data) | *Access to Facilities and Services*<br>• Availability of hospital-based cancer-related services (decreased for surgery, mammography, and endoscopy) |

| ***Author, Year*** | ***Methodology (analysis)*** | ***Geographic Scope*** | ***Period of Study*** | ***Rural Definition (threshold)*** | ***Data Sources (purpose)*** | ***Outcomes, Impacts Measured (significant finding)*** |
|---|---|---|---|---|---|---|

**Abbreviations:** ACS – American Community Survey, ACSC – Ambulatory Care Sensitive Conditions, AHA – American Hospital Association, AHRF – Area Health Resources Files, ALOS – Average Length of Stay, APRN – Advanced Practice Registered Nurse, BLS – Bureau of Labor Statistics, CBSA – Core Based Statistical Area, CHC – Community Health Center, CMS – Centers for Medicare & Medicaid Services, ECSC - Emergency Care Sensitive Conditions, ED – Emergency Department, EMS – Emergency Medical Services, FORHP – Federal Office of Rural Health Policy, FQHC – Federally Qualified Health Center, Gyn – Gynecology, HCUP – Healthcare Cost and Utilization Report, HRSA – Health Resources and Services Administration, HSA – Hospital Service Area, HSAF – Health Service Area File, IRR – Index of Relative Rurality, IRS – Internal Revenue Service, NEMSIS – National Emergency Medical Services Information System, OB – Obstetrics, OMB – Office of Management and Budget, OSHPD – Office of Statewide Health Planning and Development (California), POS – Provider of Services Current Files, PPS – Prospective Payment System, QCEW – Quarterly Census of Employment and Wages, RHC – Rural Health Clinic, RUCA – Rural-Urban Commuting Area, RUCC – Rural-Urban Continuum Codes, SID – State Inpatient Databases, UIC – Urban Influence Codes

### Detailed Description of Included Studies - Mergers

| *Author, Year* | *Methodology (analysis)* | *Geographic Scope* | *Period of Study* | *Rural Definition (threshold)* | *Data Sources (purpose)* | *Outcomes, Impacts Measured (significant finding)* |
|---|---|---|---|---|---|---|
| *Heard et al. 2022(Heard et al., 2022)* | Quantitative (retrospective cohort study) | Single institution/ system | 2015-2020 | Not reported | • Hospital system trauma registry | *Utilization*<br>• Injury severity score (increased)<br>• Hospital days<br>• Ventilator days<br>• Intensive Care Unit days<br>• Consultations with specialists (increased ortho consults)<br>*Well-being*<br>• Mortality<br>• Disposition (increased proportion of discharges to Skilled Nursing Facility, Inpatient Rehabilitation Facility) |
| *Henke et al. 2021(Henke et al., 2021a)* | Quantitative (difference in differences) | National | 2007-2018 | FORHP definition | • Irving Levin Associates data (identify merged hospitals)<br>• AHA Hospital Directory (identify merged hospitals)<br>• HCUP SID (outcome data) | *Access to Facilities and Services*<br>• Proportion of hospitals providing service lines – surgical, MSUD, maternal/fetal (decreased)<br>*Utilization*<br>• Number of hospital stays per service line (decreased) |

| *Author, Year* | *Methodology (analysis)* | *Geographic Scope* | *Period of Study* | *Rural Definition (threshold)* | *Data Sources (purpose)* | *Outcomes, Impacts Measured (significant finding)* |
|---|---|---|---|---|---|---|
| *Jiang et al. 2021(Jiang et al., 2021)* | Quantitative (case control difference in differences) | National | 2008-2019 | FORHP definition | • Irving Levin Associates data (identify merged hospitals)<br>• AHA Annual Survey (descriptive data)<br>• ACS (community characteristics)<br>• AHRF (community characteristics)<br>• HCUP SID (outcome data)<br>• AHRQ IQIs (outcome data) | *Utilization*<br>• Number of hospital stays for each IQI<br>*Well-being*<br>• Proportion of hospital stays resulting in in-hospital deaths (improved)<br>• Elective procedure complication rates |

| *Author, Year* | *Methodology (analysis)* | *Geographic Scope* | *Period of Study* | *Rural Definition (threshold)* | *Data Sources (purpose)* | *Outcomes, Impacts Measured (significant finding)* |
|---|---|---|---|---|---|---|
| *Jiang et al. 2022(Jiang et al., 2022)* | Quantitative (cohort study, survival models) | National | 2007-2019 | FORHP definition | • Irving Levin Associates data (identify merged hospitals)<br>• AHA Hospital Directory (identify merged hospitals)<br>• AHA Annual Survey (descriptive data)<br>• Sheps Center Rural Hospital Closure File (identify closed hospitals)<br>• HCUP SID (outcome data)<br>• CMS Cost Reports (outcome data)<br>• ACS (community characteristics)<br>• HCUP hospital market structure files (outcome data)<br>• KFF Medicaid Expansion Status (descriptive data) | *Well-being*<br>• Risk of closure (increased for financially stable hospitals, decreased for financially distressed hospitals)<br>*Financial*<br>• Rate of hospital financial distress (increased for new affiliations and independent hospitals)<br>• Market share (greater decrease for newly affiliated vs independent)<br>• Cost per stay (lower increase for newly affiliated vs independent)<br>*Utilization*<br>• Inpatient volume (greater decrease for newly affiliated vs independent)<br>• Length of stay<br>• Number of beds<br>• Service mix - maternal/neonatal, MSUD, surgical (increased MSUD, surgical for newly and already affiliated hospitals)<br>*Access to Facilities and Services*<br>• Distance travelled to hospital by admitted patients (greater increase for newly affiliated vs independent) |

| *Author, Year* | *Methodology (analysis)* | *Geographic Scope* | *Period of Study* | *Rural Definition (threshold)* | *Data Sources (purpose)* | *Outcomes, Impacts Measured (significant finding)* |
|---|---|---|---|---|---|---|
| *Malone et al. 2020(T. Malone et al., 2020)* | Quantitative (trend analysis) | 7 States | 2014-2016 | CBSA (non-metropolitan OR metropolitan with RUCA 4 or greater) | • Irving Levin Associates data (identify merged hospitals)<br>• HCUP SID (outcome data)<br>• CMS HCRIS (outcome data)<br>• AHA Annual Survey (descriptive data)<br>• CMS POS data (descriptive data) | *Utilization*<br>• Proportion of inpatient stays bypassing local hospital – by Major Diagnostic Category (MDC) (decreased for respiratory system discharges, mental diseases and disorders)<br>• Proportion of discharges – local, transfer, bypass<br>• Proportion of inpatient stays bypassing local hospital – by Diagnostic Related Group (DRG) |

| *Author, Year* | *Methodology (analysis)* | *Geographic Scope* | *Period of Study* | *Rural Definition (threshold)* | *Data Sources (purpose)* | *Outcomes, Impacts Measured (significant finding)* |
|---|---|---|---|---|---|---|
| *Noles et al 2015 (Noles et al., 2014, 2015)* | Quantitative (multiple regression) | National | 2005-2012 | Office of Management and Budgets (non-metropolitan county) | • Irving Levin Associates data (identify merged hospitals)<br>• CMS HCRIS (outcome data) | *Financial*<br>• Operating margin (decreased)<br>• Total profit margin<br>• Cash flow margin<br>• Earnings before interest and taxes (EBIT)<br>• Total capital expenditures<br>• Asset-to-liability ratio<br>• Proportion of equity financing<br>• Total salary expense (decreased)<br>• Salary to revenue ratio<br>*Workforce*<br>• FTEs per bed<br>• Average salary of FTE (decreased)<br>*Utilization*<br>• Proportion of revenue from outpatient services - nursery, Skilled Nursing Facility days<br>• Average acute census<br>• Annual discharges |

| ***Author, Year*** | ***Methodology (analysis)*** | ***Geographic Scope*** | ***Period of Study*** | ***Rural Definition (threshold)*** | ***Data Sources (purpose)*** | ***Outcomes, Impacts Measured (significant finding)*** |
|---|---|---|---|---|---|---|
| *O'Hanlon et al. 2019(O' Hanlon et al., 2019a)* | Quantitative (propensity score weighted controlled difference in differences) | National | 2008-2017 | FORHP definition | • AHA Annual Surveys (identify merged hospitals, descriptive data)<br>• CMS HCRIS (outcome data)<br>• CMS Hospital Compare (outcome data) | *Access to Facilities and Services*<br>• Access to technology – composite score (decreased)<br>• Availability of services – OB/Gyn, Rural Health Clinic, primary care (decreased OB and primary care)<br>*Utilization*<br>• Number of admissions<br>• Number of visits - ED, non-emergency outpatient (decreased for non-emergency outpatient)<br>*Financial*<br>• Operating margin (increased)<br>• Asset-to-liability ratio<br>• Uncompensated/unreimbursed care as a proportion of operating costs (increased, but driven by reduction in operating costs rather than increase in uncompensated care)<br>*Well-being*<br>• Patient experience – composite score<br>• 30-day readmissions - all-cause |

| ***Author, Year*** | ***Methodology (analysis)*** | ***Geographic Scope*** | ***Period of Study*** | ***Rural Definition (threshold)*** | ***Data Sources (purpose)*** | ***Outcomes, Impacts Measured (significant finding)*** |
|---|---|---|---|---|---|---|
| *Oyeka et al. 2023(Oyeka et al., 2023)* | Quantitative (propensity matched cohort) | National | 2008-2020 | RUCA 2010 (greater than 3) | • AHA Annual Surveys (identify merged hospitals, outcome data) | *Access to Facilities and Services*<br>• Proportion of hospitals offering services - 62 services<br>• Service additions (more common in hospitals that left systems)<br>• Service losses (more common in hospitals that joined systems) |
| *Williams et al. 2020(J. D. Williams, 2019; Williams Jr. et al., 2020a)* | Quantitative (difference in differences) | National | 2003-2016 | FORHP definition | • Irving Levin Associates data (identify merged hospitals)<br>• CMS HCRIS (outcome data)<br>• CMS Impact File HIPPS (outcome data)<br>• HSAF (descriptive data)<br>• AHRF (descriptive data)<br>• US Census (descriptive data) | *Financial*<br>• Total capital expenditures (increased for merged)<br>• Ability to cover debt (increased for merged)<br>• Depreciation – accumulated and expense<br>• Total revenue (decreased for merged)<br>• Total capital expenditures to revenue ratio<br>• Net patient revenue<br>*Well-being*<br>• Plant age – quartile (higher likelihood of newest quartile for merged)<br>*Utilization*<br>• Total discharges (decreased for merged)<br>• Average daily census |

| *Author, Year* | *Methodology (analysis)* | *Geographic Scope* | *Period of Study* | *Rural Definition (threshold)* | *Data Sources (purpose)* | *Outcomes, Impacts Measured (significant finding)* |
|---|---|---|---|---|---|---|
| *Williams et al. 2021(D. J. Williams et al., 2021a; J. D. Williams, 2019)* | Quantitative (difference in differences) | National | 2004-2016 | FORHP definition | • Irving Levin Associates data (identify merged hospitals)<br>• CMS HCRIS (outcome data)<br>• CMS Impact File HIPPS (outcome data)<br>• HSAF (descriptive data)<br>• AHRF (descriptive data)<br>• US Census (descriptive data) | *Financial*<br>• Inpatient charges (decreased for merged)<br>• Outpatient charges (increased for merged)<br>• Total revenue (decreased for merged)<br>• Net patient revenue (decreased for merged)<br>*Utilization*<br>• Total discharges<br>• Acute daily census |

**Abbreviations:** ACS – American Community Survey, AHA – American Hospital Association, AHRF – Area Health Resources Files, AHRQ – Agency for Healthcare Research and Quality, CBSA – Core Based Statistical Area, CMS – Centers for Medicare & Medicaid Services, ED – Emergency Department, FORHP – Federal Office of Rural Health Policy, FQHC – Federally Qualified Health Center, FTE – Full Time Equivalent, Gyn – Gynecology, HCRIS – Healthcare Provider Cost Reporting Information System, HCUP – Healthcare Cost and Utilization Report, HIPPS – Health Insurance Prospective Payment System, HSAF – Hospital Service Area Files, IQI – Inpatient Quality Indicators, MSUD – Mental and Substance Use Disorder, OB – Obstetrics, OMB – Office of Management and Budget, POS – Provider of Services Current Files, RUCA – Rural-Urban Commuting Area, RUCC – Rural-Urban Continuum Codes, SID – State Inpatient Databases.

### Health System Ecologies Impact Matrices

Impact Matrix for Rural Hospital Closures

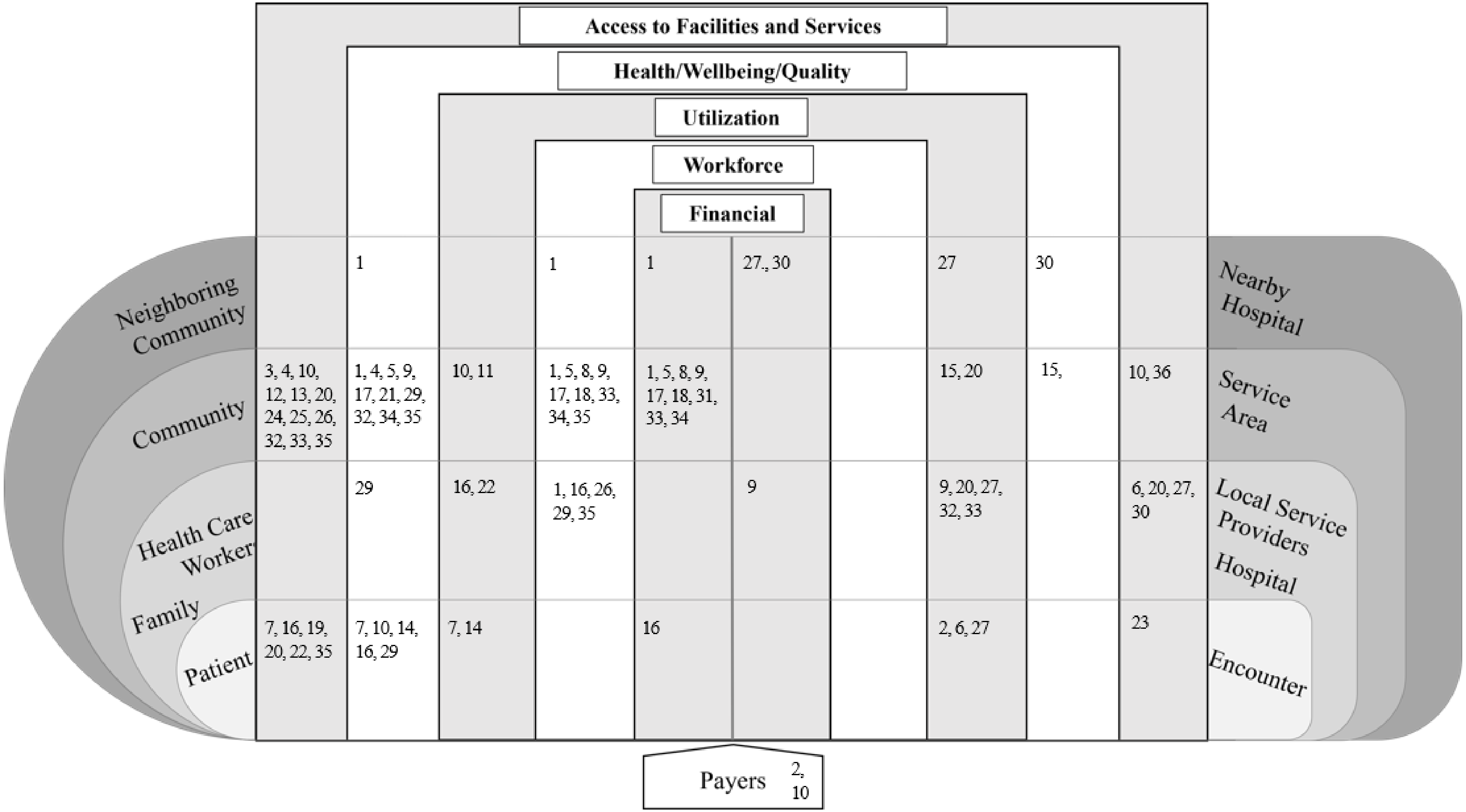

*Index of Included Studies of Rural Hospital Closures*

| | |
|---|---|
| 1. (Alexander & Richards, 2023)<br>2. (Andreyeva et al., 2022)<br>3. (Bell et al., 2023)<br>4. (Capital Link, 2022)<br>5. (Chatterjee et al., 2022)<br>6. (Chaudhary et al., 2019)<br>7. (Durrance et al., 2024)<br>8. (Edmiston, 2019)<br>9. (Eilrich et al., 2015)<br>10. (US Government Accountability Office, 2020)<br>11. (Gelbaugh & Advisory Board, 2021)<br>12. (Germack et al., 2019)<br>13. (Germack et al., 2021)<br>14. (Gujral & Basu, 2019)<br>15. (Khushalani et al., 2022)<br>16. (Letheren et al., 2024)<br>17. (T. L. Malone et al., 2022)<br>18. (Manlove & Whitacre, 2017) | 19. (McCarthy et al., 2021)<br>20. (Medicare Payment Advisory Commission (MEDPAC), 2021)<br>21. (Merrell, 2019)<br>22. (Mike, 2020)<br>23. (Miller et al., 2020)<br>24. (Miller et al., 2021)<br>25. (Mills, 2022)<br>26. (Mobley et al., 2020)<br>27. (Nikpay et al., 2021)<br>28. (Ramedani et al., 2022)<br>29. (J. G. Smith et al., 2022)<br>30. (T. B. Smith et al., 2022)<br>31. (Song & Saghafian, 2019)<br>32. (Tennessee Health Care Campaign, 2021)<br>33. (Thomas et al., 2015)<br>34. (Vogler, 2020)<br>35. (Wishner et al., 2016)<br>36. (Zahnd et al., 2023) |

Impact Matrix for Rural Hospital Mergers

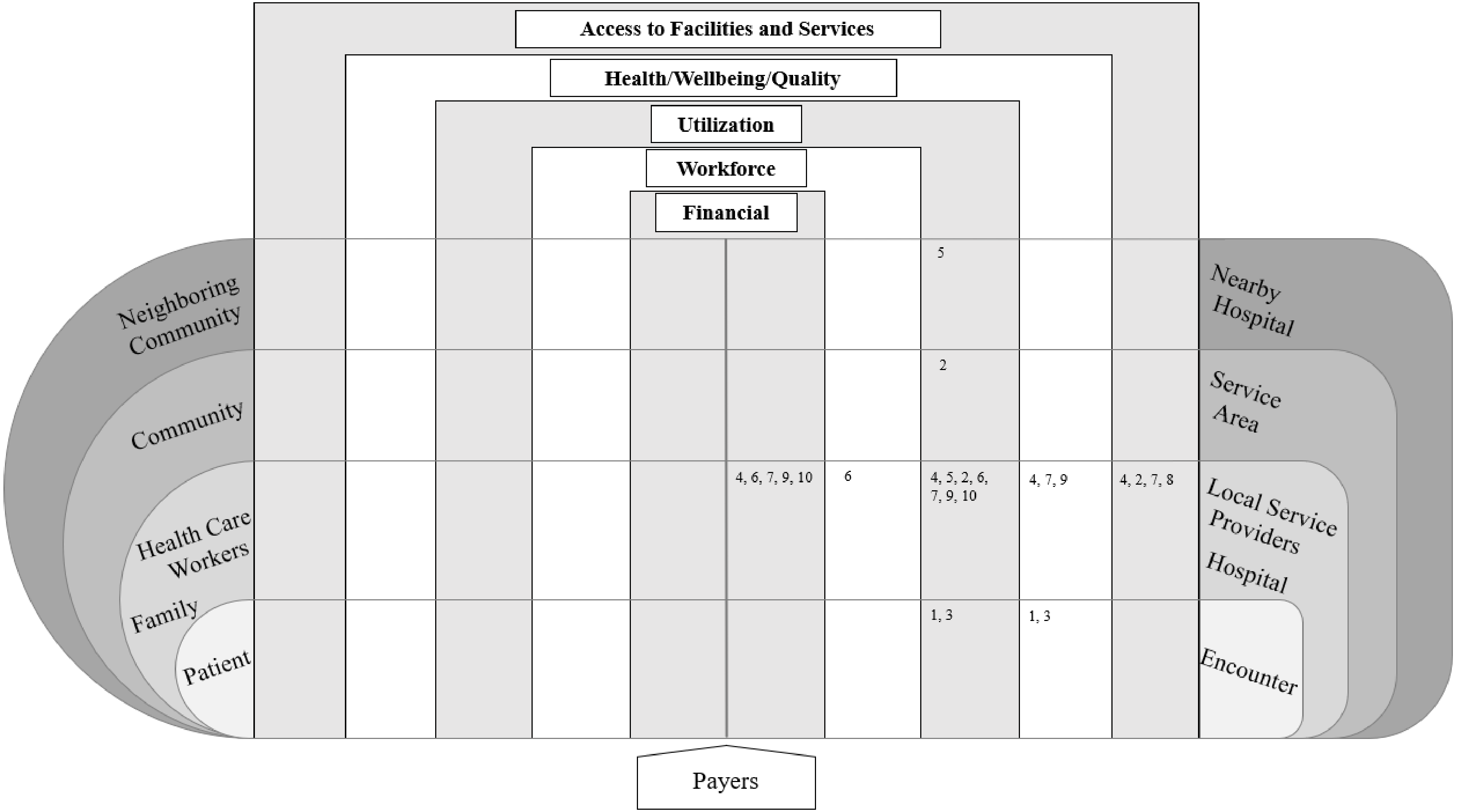

*Index of Included Studies of Rural Hospital Closures*
